# Respiratory Fungal Communities are Associated with Systemic Inflammation and Predict Survival in Patients with Acute Respiratory Failure

**DOI:** 10.1101/2023.05.11.23289861

**Authors:** Noel Britton, Haopu Yang, Adam Fitch, Kelvin Li, Khaled Seyed, Rui Guo, Shulin Qin, Yingze Zhang, William Bain, Faraaz Shah, Partha Biswas, Wonseok Choi, Malcolm Finkelman, Yonglong Zhang, Catherine L. Haggerty, Panayiotis V. Benos, Maria M. Brooks, Bryan J. McVerry, Barbara Methe, Georgios D. Kitsios, Alison Morris

**Author notes:** <u>Corresponding Authors:</u> Alison Morris, MD, MS, Professor of Medicine, Chief, Pulmonary, Allergy, Critical Care, and Sleep Medicine Division, UPMC Chair of Translational Pulmonary and Critical Care Medicine, Director, University of Pittsburgh Center for Medicine and the Microbiome, Georgios Kitsios, MD PhD, Assistant Professor of Medicine, Pulmonary, Allergy, Critical Care, and Sleep Medicine Division, University of Pittsburgh. co-senior authors (Georgios D. Kitsios, Alison Morris).

## Abstract

**Rationale:** Disruption of respiratory bacterial communities predicts poor clinical outcomes in critical illness; however, the role of respiratory fungal communities (mycobiome) is poorly understood.

**Objectives:** We investigated whether mycobiota variation in the respiratory tract is associated with host-response and clinical outcomes in critically ill patients.

**Methods:** To characterize the upper and lower respiratory tract mycobiota, we performed rRNA gene sequencing (internal transcribed spacer) of oral swabs and endotracheal aspirates (ETA) from 316 mechanically-ventilated patients. We examined associations of mycobiome profiles (diversity and composition) with clinical variables, host-response biomarkers, and outcomes.

**Measurements and Main Results:** ETA samples with >50% relative abundance for *C. albicans* (51%) were associated with elevated plasma IL-8 and pentraxin-3 (p=0.05), longer time-to-liberation from mechanical ventilation (p=0.04) and worse 30-day survival (adjusted hazards ratio (adjHR): 1.96 [1.04-3.81], p=0.05). Using unsupervised clustering, we derived two clusters in ETA samples, with Cluster 2 (39%) showing lower alpha diversity (p<0.001) and higher abundance of *C. albicans* (p<0.001). Cluster 2 was significantly associated with the prognostically adverse hyperinflammatory subphenotype (odds ratio 2.07 [1.03-4.18], p=0.04) and predicted worse survival (adjHR: 1.81 [1.03-3.19], p=0.03). *C. albicans* abundance in oral swabs was also associated with the hyper-inflammatory subphenotype and mortality.

**Conclusions:** Variation in respiratory mycobiota was significantly associated with systemic inflammation and clinical outcomes. *C. albicans* abundance emerged as a negative predictor in both the upper and lower respiratory tract. The lung mycobiome may play an important role in the biological and clinical heterogeneity among critically ill patients and represent a potential therapeutic target for lung injury in critical illness.

## INTRODUCTION

Disruption of the bacterial communities in the intubated respiratory tract has been linked to more severe illness, prolonged time on mechanical ventilation, and worse survival in patients with acute respiratory failure, as evidenced by several studies. (1–7) In contrast, the role of respiratory fungi (i.e., the mycobiome) in acute respiratory failure remains understudied. Recent research suggests that mycobiota may play a significant role in critical illness. (8–12) Although yeast organisms are often detected in respiratory specimen cultures, they are typically considered to represent airway colonization rather than infection. (13–21) Nonetheless, fungal colonization can trigger regional immune responses and mediate lung damage in acute and chronic illness outside conventionally defined invasive fungal infection. (13–15, 21–28) In fact, *Candida* spp. airway colonization has been associated with worse outcomes in critically ill patients. (14, 15, 21, 29, 30) Most studies of *Candida* colonization have used conventional microbial culture-based methods, which are limited in sensitivity for certain fastidious fungal species, lack the ability to profile the fungal community comprehensively, have limited resolution in species identification, and provide narrow insight into the interactions of fungal and bacterial communities. To address these knowledge gaps, we characterized the upper and lower respiratory tract mycobiome in mechanically ventilated critically ill patients and examined the associations between lung mycobiome features, the systemic host response, and clinical outcomes. We hypothesized that mycobiome dominance and decreased ecological diversity are associated with worse clinical outcomes.

## METHODS

### Detailed methods are provided in the Supplement

#### Patient Enrollment and Clinical Characteristics

We enrolled adult patients with acute respiratory failure within 72 hours of initiation of mechanical ventilation. We sampled upper and lower respiratory tract mycobiota with a posterior oropharyngeal (oral) swab and an endotracheal aspirate (ETA), respectively. (1) We collected blood samples for measurement of host-response biomarkers in plasma. We extracted clinical data from the electronic medical record and utilized clinical microbiologic results obtained within 48hrs of research biospecimens. (31) We considered yeast growth in cultures as fungal colonization, mirroring the interpretation of yeast detection in routine clinical practice. (31) We followed patients prospectively for time-to-liberation from mechanical ventilation and 30-day survival. (32)

#### Laboratory Analyses

In plasma, we measured ten host response biomarkers and the fungal cell wall constituent (1→3)-β-D-Glucan (BDG). (1, 31) We extracted microbial DNA from ETA samples and oral swabs, which we amplified by PCR targeting the internal transcribed spacer (ITS) and then sequenced on the Illumina Miseq platform. (31) From derived ITS sequences, we applied a custom pipeline for amplicon sequence variant (ASV) classification (31, 33, 34) and excluded samples that generated fewer than 100 reads.

#### Data processing and statistical analyses

We compared clinical variables between patients with samples excluded due to low sequencing reads and those included using Wilcoxon rank sum tests for continuous variables and Fisher’s exact test for categorical variables, and tested differences in fungal load (number of sequencing reads) between clinical samples and experimental controls with Wilcoxon tests. We performed ecological analyses of alpha (Shannon index) and beta (Manhattan distances with permutational analysis of variance [PERMANOVA]) diversity and visualized beta diversity using principal coordinates analyses (PCoA) plots. Relative abundance was calculated at the fungal species level; mycobiome dominance was defined as relative abundance of a species greater than 50%. Samples were categorized as “monofungal” when only a single species was identified. Biomarker values were log-transformed for analyses. We assigned patients to a hyper-vs. hypo-inflammatory subphenotype based on a validated, parsimonious regression model utilizing four plasma biomarker values (soluble Tumor Necrosis Factor Recptor-1 [sTNFR1], angiopoietin-2 [Ang-2], procalcitonin, and bicarbonate). (35)

Using a supervised approach, we analyzed specific features of fungal communities individually: alpha (Shannon index) and beta diversity, fungal load, relative abundance, and mycobiome dominance (single species relative abundance ≥ 50%). Multivariate linear regression models were created to determine associations with features of fungal communities with plasma biomarker levels. We constructed Cox proportional hazards and Fine-Gray (treating death as a competing risk) models for 30-day survival and time-to-liberation from mechanical ventilation, respectively. Adjusted models included covariates selected based on clinical judgment and backward stepwise regression, including age, history of pulmonary fibrosis, and sequential organ failure assessment (SOFA) scores. (36) We used unsupervised Dirichlet Multinomial Models (DMM) with Laplace approximations to agnostically examine for distinct clusters of fungal composition. (1, 37) From our previously published work, we utilized 16S rRNA gene sequencing data and DMM assignment to clusters of bacterial profiles from the same cohort of patients for comparisons between fungal and bacterial communities. (1) An unsupervised approach was used to evaluate clinical and biomarker differences between the derived DMM clusters. All analyses were conducted in R (v.4.1.2).

## RESULTS

### Sequencing yield and quality control

We enrolled 316 mechanically-ventilated patients who contributed 538 airway samples at the time of enrollment (222 oral swabs and 316 ETAs) (**Supplementary Figure 1**). We excluded 78 ETA samples and 20 oral samples due to poor sequencing yield (<100 reads) (**Supplementary Tables 2 and 3**). Compared to participants with included samples (n=238), those with ETA samples excluded (n=78) for low reads were less likely to have positive fungal respiratory cultures (8.3% vs. 21.7%, p=0.02) (**Supplementary Table 2**).

### Upper and lower respiratory tract fungal communities exhibit substantial heterogeneity

Clinical samples from ICU patients had a significantly higher number of ITS reads compared to the 96 procedural control samples (**Figure 1, Supplementary Table 5**). Microbial profiles from both oral swabs and ETAs exhibited substantial heterogeneity. ETA samples had markedly fewer fungal reads (median 3228, interquartile range [109.5, 27245]) compared to oral samples (median 20828.5 [2367.5, 49991], p<0.001) (**Figure 1A, Supplementary Table 5**). Alpha diversity was not significantly different between ETA and oral samples (p=0.15) and ranged from Shannon index of zero (effectively monofungal communities) to > 2.0 (**Figure 1B, Supplementary Table 6**). The proportion of monofungal communities differed between oral (n=37, [18.3%]) and ETA (n=68 [28.6%]) samples (p=0.01) (**Supplementary Figure 2**). By beta diversity comparisons, individual samples were distributed over wide spaces of PCoA plots indicating wide variation in fungal species composition (**Figure 1C**) (p<0.001). We identified 99 different fungal species with greater than 0.005 relative abundance. *Candida albicans* (*C. albicans*) was the most abundant in oral and ETA samples, followed by *C. dubliniensis, C. tropicalis*, and *C. parapsilosis*. The most common non-Candida fungal genera included *Cladosporium, Meyerozyma, Pneumocystis*, and *Saccharomyces* spp. *C. albicans* relative abundance in oral and ETA samples was significantly correlated (r=0.62, p<0.001, Supplementary Figure S3).

**Figure 1:**
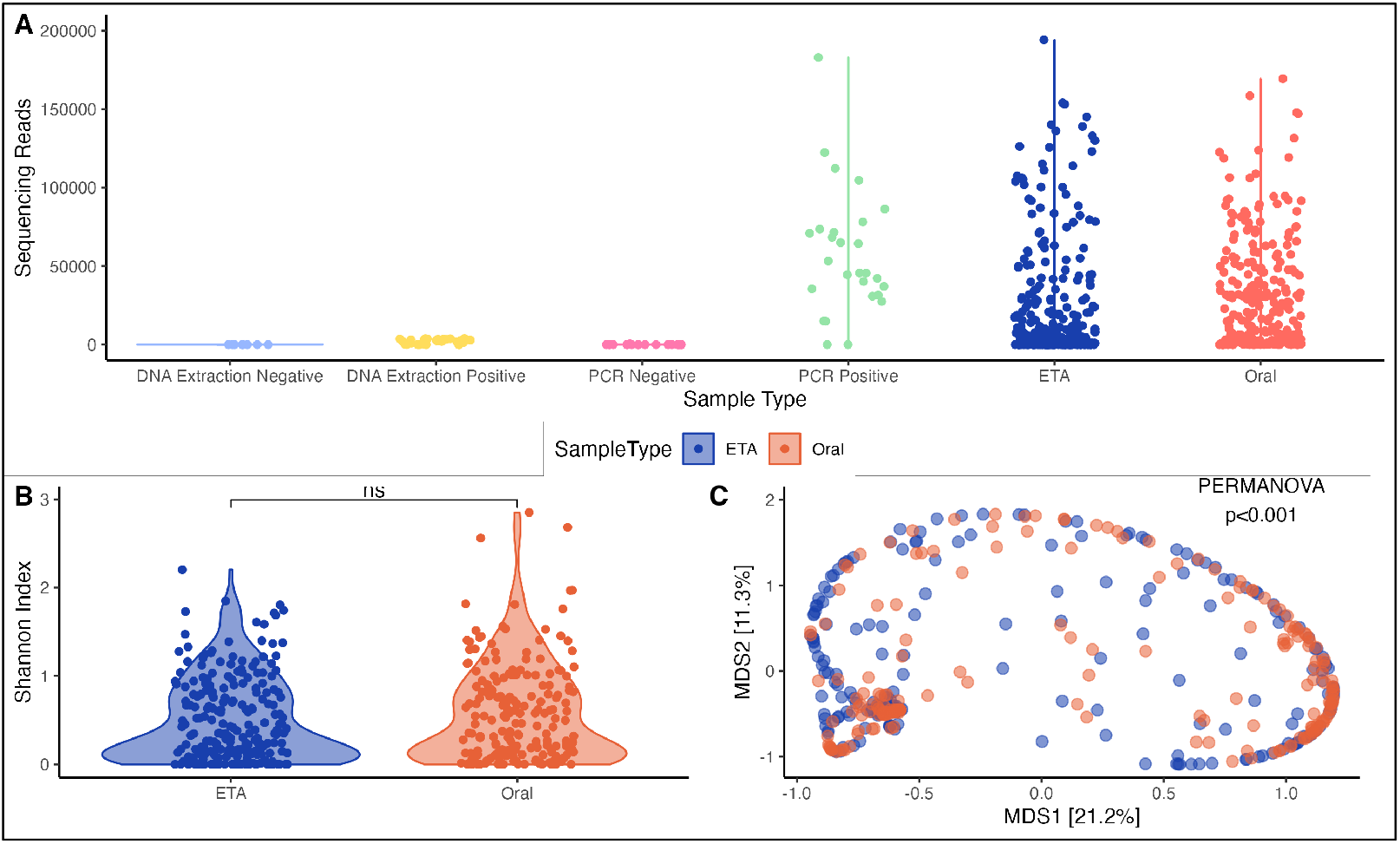
Ecological analyses of fungal load, alpha and beta diversity for ETA and oral samples. (A) Number of high-quality ITS reads per sample are shown on the y-axis. Clinical samples from both ETA and oral samples had much higher ITS reads than experimental control samples (p<0.001). We excluded 81 ETA and 21 oral samples due to poor sequencing yield (<100 reads; not shown). ETA samples had a greater number of fungal sequencing reads compared to oral samples. (B) The Shannon index was not significantly different between ETA and oral samples. (C) Principal coordinates analyses for beta-diversity comparisons (Manhattan distances) with PERMANOVA for oral and ETA samples. ETA= endotracheal aspirate; PERMANOVA = permutational ANOVA; MDS = multidimensional scaling axes; ns: p > 0.05; ^*^ p ≤ 0.05; ^**^ p ≤ 0.01; ^***^ p ≤0.001; ^****^ p ≤0.0001

### Cohort description

Of the 238 participants (mean age 60 years, 56.3% male) with samples included in the analysis, 25.2% were diagnosed with ARDS, 85.7% were receiving systemic antibiotics at the time of sampling in the ICU, and 21.6% were classified to the hyperinflammatory subphenotype (**Table 1**).

**Table 1:**
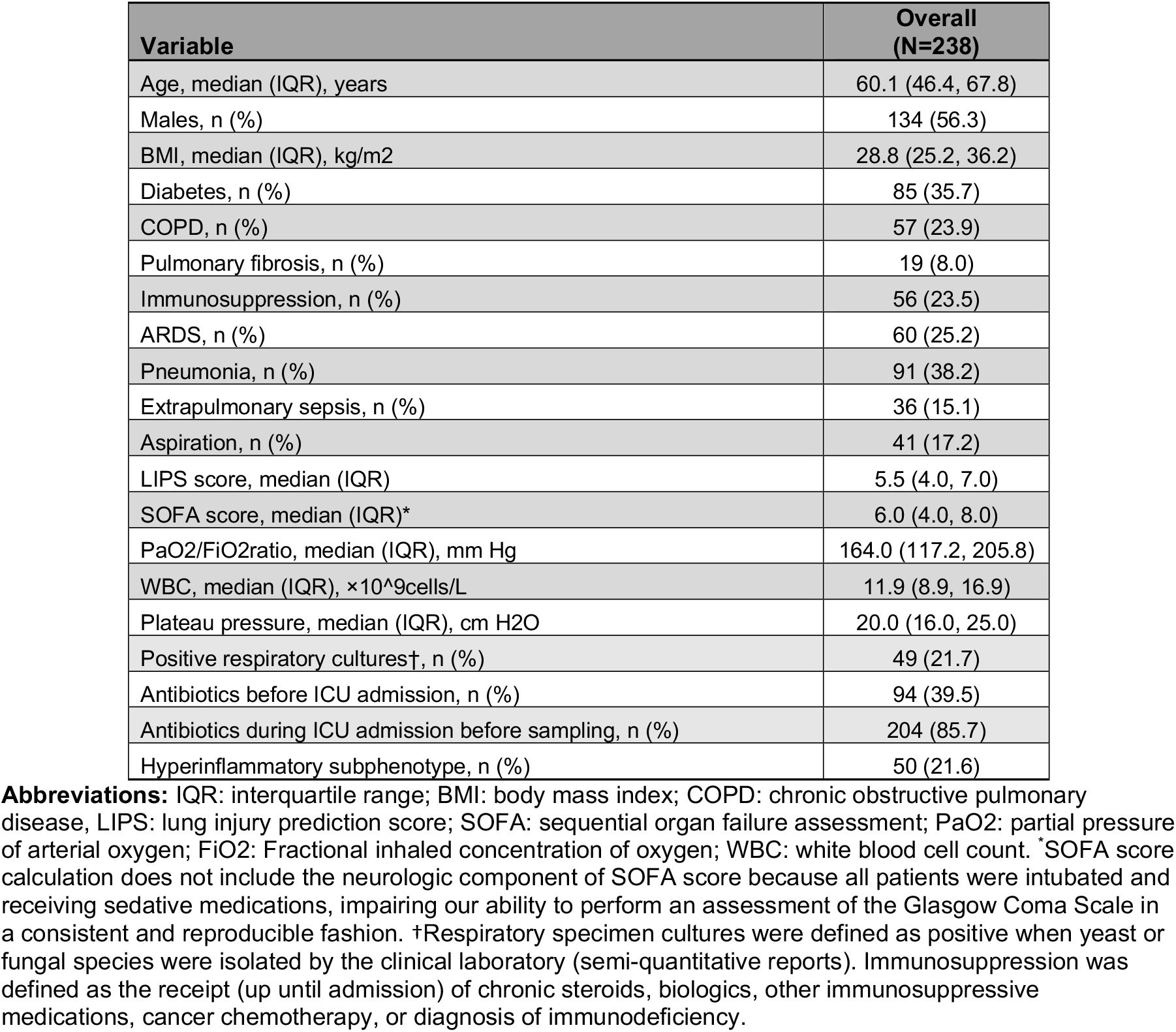
Baseline characteristics and clinical outcomes of patients included in analysis. Data are presented as median (with interquartile ranges) for continuous variables and N (%) for categorical variables.

| Variable | Overall (N=238) |
| --- | --- |
| Age, median (IQR), years | 60.1 (46.4, 67.8) |
| Males, n (%) | 134 (56.3) |
| BMI, median (IQR), kg/m<sup>2</sup> | 28.8 (25.2, 36.2) |
| Diabetes, n (%) | 85 (35.7) |
| COPD, n (%) | 57 (23.9) |
| Pulmonary fibrosis, n (%) | 19 (8.0) |
| Immunosuppression, n (%) | 56 (23.5) |
| ARDS, n (%) | 60 (25.2) |
| Pneumonia, n (%) | 91 (38.2) |
| Extrapulmonary sepsis, n (%) | 36 (15.1) |
| Aspiration, n (%) | 41 (17.2) |
| LIPS score, median (IQR) | 5.5 (4.0, 7.0) |
| SOFA score, median (IQR)* | 6.0 (4.0, 8.0) |
| PaO<sub>2</sub>/FiO<sub>2</sub> ratio, median (IQR), mm Hg | 164.0 (117.2, 205.8) |
| WBC, median (IQR), ×10<sup>9</sup> cells/L | 11.9 (8.9, 16.9) |
| Plateau pressure, median (IQR), cm H<sub>2</sub>O | 20.0 (16.0, 25.0) |
| Positive respiratory cultures†, n (%) | 49 (21.7) |
| Antibiotics before ICU admission, n (%) | 94 (39.5) |
| Antibiotics during ICU admission before sampling, n (%) | 204 (85.7) |
| Hyperinflammatory subphenotype, n (%) | 50 (21.6) |

**Abbreviations:** IQR: interquartile range; BMI: body mass index; COPD: chronic obstructive pulmonary disease, LIPS: lung injury prediction score; SOFA: sequential organ failure assessment; PaO<sub>2</sub>: partial pressure of arterial oxygen; FiO<sub>2</sub>: Fractional inhaled concentration of oxygen; WBC: white blood cell count. \*SOFA score calculation does not include the neurologic component of SOFA score because all patients were intubated and receiving sedative medications, impairing our ability to perform an assessment of the Glasgow Coma Scale in a consistent and reproducible fashion. †Respiratory specimen cultures were defined as positive when yeast or fungal species were isolated by the clinical laboratory (semi-quantitative reports). Immunosuppression was defined as the receipt (up until admission) of chronic steroids, biologics, other immunosuppressive medications, cancer chemotherapy, or diagnosis of immunodeficiency.

### ETA mycobiome features are associated with host-response

In multivariate-adjusted linear models, we examined for associations between plasma biomarkers and ETA fungal community features (fungal load, Shannon index, and *C. albicans* relative abundance). Fungal load was significantly associated with higher plasma IL-8 (p=0.05) after adjusting for covariates. *C. albicans* relative abundance was also significantly associated with increased plasma levels of ANG-2 (p=0.01) and BDG> 60 pg/mL (0.05). We arbitrarily define a sample to be dominated by a single fungal taxon if the relative abundance of which surpasses 50%. We found that *C. albicans* dominated (relative abundance ≥ 50%) 51% of ETA samples, and such patients were more likely to have been treated with immunosuppression (p=0.05) and have a respiratory culture positive for yeast (p=0.02). *C. albicans-*dominated samples were associated with elevated levels of plasma IL-8 (p=0.05), ANG-2 (p=0.03), pentraxin-3 (p=0.03), and BDG >60 pg/mL.

### Supervised analyses reveal *C. albicans* relative abundance as a predictor of outcome

There were no significant differences in 30-day mortality [survivors vs. non-survivors] by fungal load, Shannon index, beta diversity, the relative abundance of *C. albicans*, mycobiome dominance, or respiratory fungal culture result in ETA sample. We saw no change in these results in a sensitivity analysis excluding the six patients with positive fungal culture indicative of invasive fungal infection. In Cox proportional hazards models, greater *C. albicans* relative abundance was associated with worse 30-day survival (Adj. HR 1.96, CI 1.24-3.81, p=0.05) and longer time to liberation from mechanical ventilation (Adj. HR 0.67, CI: 0.46-0.98, p=0.04).

### Lower respiratory communities comprise distinct compositional clusters that are prognostic of outcome

Demonstrating heterogeneous ETA and oral sample fungal composition in terms of fungal load, alpha, and beta diversity, we pursued an unsupervised analysis to better understand the fungal profile heterogeneity. With the DMM method, Laplace approximation of model fitting showed that two clusters offered the best fit in ETA samples. Cluster 2 was characterized by higher fungal load (**Figure 2A**), lower Shannon index (p<0.001) (**Figure 2B**), significantly different taxonomic composition (adjusted PERMANOVA p<0.001 for beta-diversity differences, **Figure 2C** and **Figure 3A**) as well as higher relative abundance of *C. albicans* compared to Cluster 1 (**Figure 3B**). We examined the association of fungal DMM clusters with previously published bacterial DMM clusters from the same cohort. Bacterial DMM clusters 1 and 3 had high abundance of typical members of the respiratory microbiome (i.e., oral-origin bacteria, such as *Streptococcus, Prevotella*, and *Veillonella*). In contrast, bacterial DMM cluster 2 had lower Shannon diversity and was associated with significantly worst outcomes compared to clusters 1 and 3. There was no significant association between bacterial and fungal DMM clusters (Fisher’s Exact test p=0.21) (**Figure 4**).

**Figure 2:**
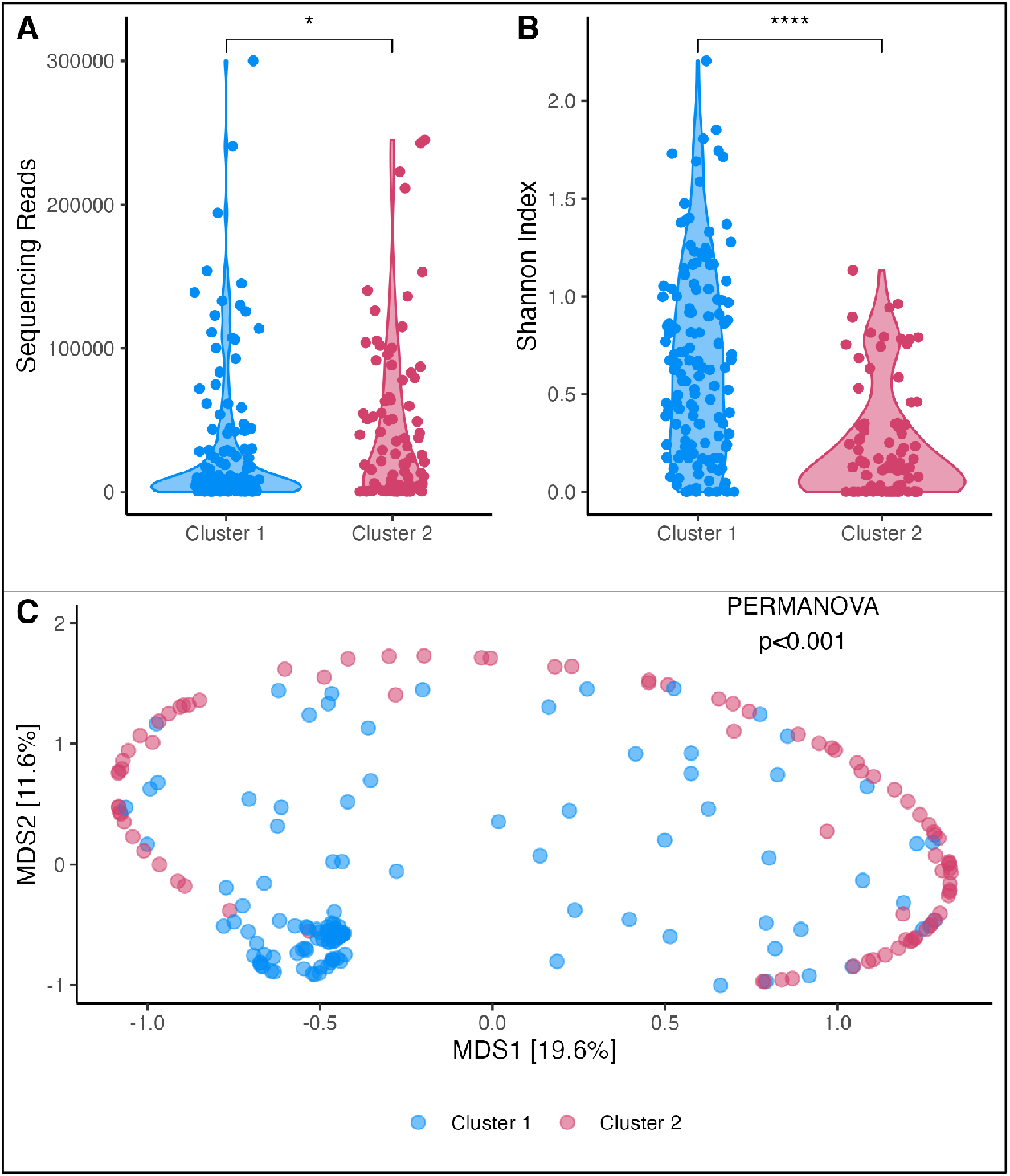
Dirichlet-multinomial-model clustering of ETA communities reveals a distinct cluster marked by low alpha diversity. (A) Number of high-quality ITS reads per sample are shown on the y-axis. No significant difference in the number of sequencing reads was observed between clusters. (B) Cluster 2 samples had a lower Shannon index than Cluster 1 (p<0.001). (C) Principal coordinates analyses for β-diversity comparisons (Manhattan distances) with PERMANOVA for all samples included and stratified by clusters. DMM= Dirichlet multinomial model; ETA= endotracheal aspirate; ITS= internal transcribed spacer; PERMANOVA = permutational ANOVA; MDS = multidimensional scaling axes; ns: p > 0.05; ^*^ p ≤ 0.05; ^**^ p ≤0.01; ^***^ p ≤0.001; ^****^ p ≤0.0001

**Figure 3:**
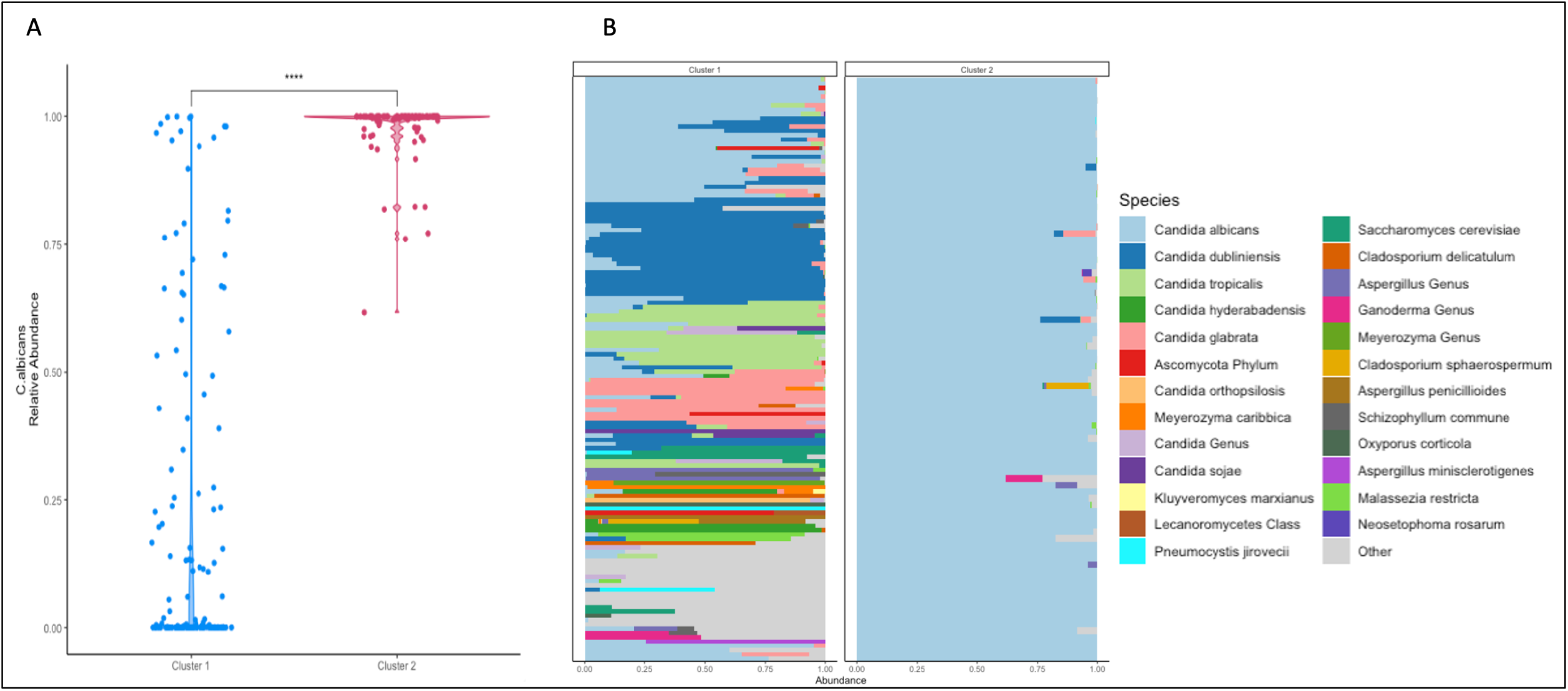
Taxonomic compositional differences between Dirichlet Multinomial Model clusters. (A) The relative abundance of *C. albicans* stratified between clusters. Cluster 2 had higher relative abundance of *C*.*albicans* than Cluster 1. (B) Taxonomic bar plots for individual endotracheal aspirate samples, stratified by Dirichlet Multinomial Models clusters. Taxonomic composition is shown as stacked bar graphs, with each bar representing the fungal community of each cluster, with taxa colored individually and widths of component bars corresponding to the relative abundance of each species for the top 25 species. ns: p > 0.05; ^*^ p ≤0.05; ^**^ p ≤ 0.01; ^***^ p ≤ 0.001; ^****^ p ≤0.000

**Figure 4:**
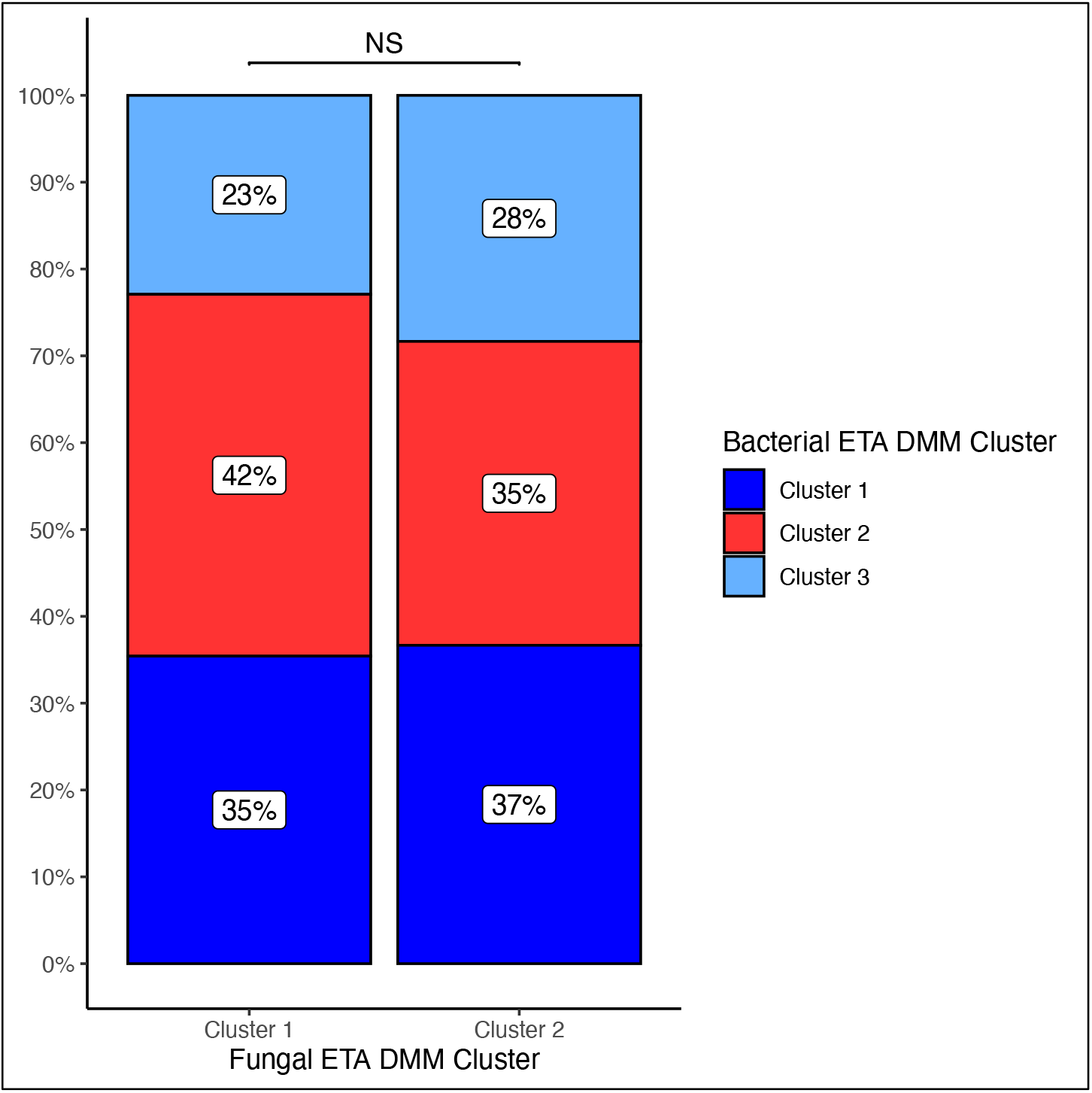
Bacterial Dirichlet Multinomial Model cluster assignment by Fungal Dirichlet Multinomial Model cluster assignment for endotracheal aspirates. Data are presented as n (% of fungal DMM cluster). Fisher’s Exact Test, p=0.64.

ETA Cluster 2 patients (i.e., the *C. albicans*-enriched cluster with low alpha diversity) were more likely to have a history of immunosuppressive treatment (p=0.02) and were more often diagnosed with extrapulmonary sepsis (p=0.03) compared to Cluster 1. In multivariate linear models, Cluster 2 membership was significantly associated with higher IL-8 (adjusted p=0.03), pentraxin-3 (adjusted p=0.04), and ANG-2 (adjusted p=0.01)(**Supplementary Table 8**). Furthermore, Cluster 2 subjects had higher odds of belonging to the hyperinflammatory subphenotype (OR 2.07, CI: 1.03-4.18, p=0.04).

Consistent with the associations with worse inflammatory profiles, ETA Cluster 2 membership predicted worse 30-day survival (Adj. HR: 1.81, CI 1.03-3.19, p=0.03) (**Figure 5A**) as well as longer time to liberation from mechanical ventilation (Adj. HR: 0.38, CI: 0.23-0.64, p<0.001) (**Figure 5B**) compared to Cluster 1 subjects.

**Figure 5:**
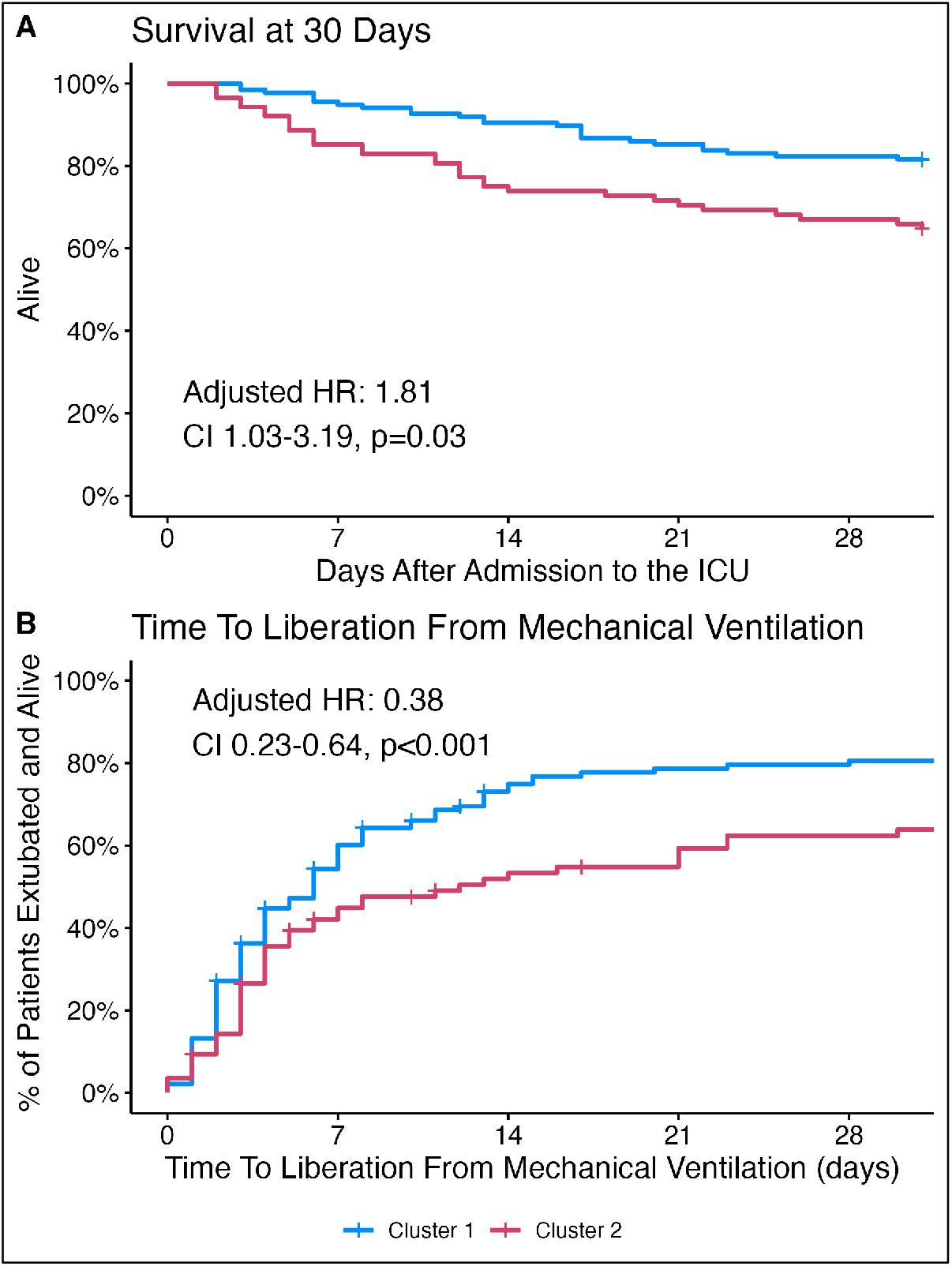
Patients with ETA samples belonging to Cluster 2 had worse 30-day survival and longer time to liberation from mechanical ventilation. (A and B) Kaplan-Meier curves for 30-day survival and time to liberation stratified by Dirichlet-multinomial-model (DMM) clusters for endotracheal aspirates (ETAs). P values were derived from the log-rank test, and hazard ratios (HRs) with corresponding 95% confidence intervals were derived from the Cox proportional hazard model adjusted for age, SOFA score, history of pulmonary fibrosis, and positive respiratory fungal culture. HR = hazards ratio.

### Oral mycobiota are associated with host response and outcome

Among the different oral community features (load, diversity, species abundance, and *C. albicans* dominance), we found that *C. albicans* dominance was the only oral mycobiome variable significantly associated with host response. Oral *C. albicans* dominance was significantly associated with elevated plasma levels of IL-8 (p<0.01), sTNFR-1 (p=0.03), pentraxin-3 (p<0.01), RAGE (p=0.03), ANG-2 (p<0.01), as well as with the hyperinflammatory subphenotype (odds ratio 2.39 [1.92, 3.76], p<0.01). Non-survivors had higher oral relative abundance of *C. albicans* compared to survivors (97.4% vs. 51.8%, p=0.006). In time-to-event analyses, *C*.*albicans* dominance was associated with prolonged time-to-liberation from mechanical ventilation (AdjHR= 0.40 [0.22, 0.79], p=0.002).

With DMM methodology, we identified two distinct clusters in oral samples. Oral Cluster 2 subjects had lower Shannon index (p<0.001), significant differences in overall composition (PERMANOVA p<0.001), and higher relative abundance of *C. albicans* (p=0.02) compared to Cluster 1 subjects. Oral Cluster 2 subjects showed higher levels of plasma BDG (p=0.04) and interleukin-10 (IL-10) (p=0.03) compared to Cluster 1 but had no differences in clinical outcomes. We examined for associations between oral and ETA DMM clusters but found no significant relationship (Fisher’s Exact Test, p=0.76).

## DISCUSSION

Our study examined the relationship between variations in respiratory tract mycobiota and clinical outcomes in a diverse group of patients with acute respiratory failure who were mechanically ventilated. We found significant heterogeneity in fungal DNA load, alpha diversity, and composition. Notably, *Candida* was the most abundant fungal genus, with *C. albicans* accounting for more than 50% relative abundance in most ETA samples. We identified features of fungal communities that were significantly associated with host response and detectable levels of BDG in the plasma. We observed correlations between *C. albicans* and plasma inflammatory biomarkers and worse survival outcomes. These observations align with previous literature demonstrating that patients with elevated BDG had higher rates of Candida spp. positive respiratory cultures and were much more likely to die by day 28 than those with BDG < 60 pg/mL. (24)

Our unsupervised analysis of ETA mycobiota identified a low diversity, *C. albicans*-enriched cluster that was significantly associated with a hyperinflammatory subphenotype and poor prognosis. Fungal airway colonization is common in ICU patients, with nearly 60% incidence estimated by culture-dependent methods. However, true fungal infections are infrequent. *Candida* spp. or unspeciated yeast detection in lower respiratory tract cultures, such as ETA or bronchoalveolar lavage fluid, poses a clinical challenge as this finding is routinely considered colonization. Still, Candida airway colonization has been linked to extended mechanical ventilation, longer ICU stays, and increased ventilator-associated pneumonia risk. (38) A small-scale randomized clinical trial demonstrated no advantage in targeting such colonization with antifungal treatment, suggesting that empiric antifungal therapy for *Candida* airway colonization is not indicated. (16) Nevertheless, the negative prognostic associations of colonization raise important questions regarding the biological consequences of abundant fungal communities in the lower respiratory tract.

The role of *Candida* airway colonization in experimental lung injury is still unclear. Previous research has indicated that *C*.*albicans* colonization triggers neutrophil infiltration in the lungs of mice (39, 40). Additionally, *C*.*albicans* can penetrate epithelial cells by direct interaction between hyphae and epithelial cells, resulting in hyphal-induced apoptosis and necrosis of epithelial cells. (41) These pathogenic mechanisms allow *C*.*albicans* to breach the endothelial and epithelial barriers, leading to increased pulmonary capillary permeability. (42) This increase in permeability leads to pulmonary edema, which is a defining characteristic of ARDS. (42, 43) In animal models, exposure to *C*.*albicans* has been shown to increase pulmonary edema. (44) These preclinical findings support our conclusion that *C*.*albicans* colonization is linked to worse clinical outcomes in mechanically ventilated critically ill patients.

Our approach to profiling the fungal communities in the upper and lower respiratory tract with culture-independent methods revealed important associations between fungal community features and the systemic host response, as well as adverse clinical outcomes. Our supervised and unsupervised analyses underscored a pattern of fungal dysbiosis, characterized by low alpha diversity and *C. albicans* enrichment, as a robust predictor of systemic inflammatory profiles and adverse outcomes, even after accounting for confounders. Notably, the fungal clusters we identified were distinct from the bacterial composition clusters we described previously in a subset of patients from this cohort. This indicates that fungal dysbiosis did not mirror the bacterial composition differences we previously reported. Our previous research demonstrated significant associations between host-response subphenotypes and ETA bacterial community clusters, (1) as well as the bacterial cell-free DNA load in plasma. (47) Although precision medicine approaches in critical illness have focused on host-response subphenotyping, the biological determinants of inflammatory subphenotypes remain poorly understood. Our findings suggest that the lung mycobiome is a previously overlooked source of biological heterogeneity during acute respiratory failure. Moreover, the statistical independence between fungal and bacterial cluster membership highlights the fact that prior bacteriome analyses have only provided a partial understanding of inter-individual microbiota variation.

Our analysis of both upper and lower respiratory tract samples provided similar insights into the composition of fungal communities and their prognostic effects on host response and outcome. Specifically, we found significant correlations between *C. albicans* abundance in oral and ETA samples, and our unsupervised clustering analyses revealed similar representations of fungal dysbiosis, characterized by low diversity communities and increased *C. albicans* abundance, which predicted adverse outcomes. Despite the high incidence of *Candida* colonization in the oropharynx of ICU patients, routine examination for fungal colonization is not typically performed in clinical practice. Clinicians often rely on macroscopic diagnosis of oropharyngeal candidiasis (i.e., observing “thrush” in the oropharynx) and treat with topical or systemic antifungals, even without microbiologic confirmation of mucosal candidiasis. Our culture-independent analysis of oropharyngeal swabs and their similarities with lower respiratory tract fungal communities provides further evidence of mouth-lung immigration shaping the microbial topography of the respiratory tract. (48, 49) With the shift towards non-invasive respiratory support in acute respiratory failure, sampling the oropharynx for microbiota analyses may offer a useful and accessible means of evaluating lower respiratory tract processes in non-intubated patients.

Our study has several limitations. First, we enrolled participants from ICUs at a single tertiary center, which may limit the generalizability of our results to broader populations. Furthermore, we conducted analyses with non-invasive samples at a single time-point, which did not allow us to examine longitudinal changes in the respiratory mycobiome. While we adjusted for clinical confounders, residual confounding may still account for some of the observed mycobiota-outcome associations. Moreover, ITS rRNA gene sequencing has limited resolution in accurately representing all fungal species in a clinical sample. We were unable to infer viability or virulence factors of specific fungi. Nonetheless, we were able to detect the clinically most relevant fungi, including *Candida* and *Aspergillus* species, and found significant associations between *Candida* spp. abundance with detectable yeast growth on clinical cultures. It is worth noting that several ETA and oral samples had low sequencing yield despite the sensitivity of PCR amplification and ITS sequencing. We found slight differences in patient characteristics between excluded and included samples, most notably the difference in clinical culture positivity for yeast growth, indicating that excluded samples had low or absent fungal signal. We used ETA samples instead of BAL due to practical reasons and the advantage of non-invasive sampling for achieving a large sample size compared to bronchoscopy-based studies. Our sampling strategy did not allow us to study regional variability of fungal communities in the lung, but instead we drew inferences on global fungal communities in the lower respiratory tract in a sample that is easily applied in the clinical setting.

In summary, we observed that reduced diversity in fungal communities in the respiratory tract of critically ill patients is associated with a hyperinflammatory subphenotype, higher mortality and longer periods of mechanical ventilation. We discovered that patients with enrichment for *C. albicans* in both upper and lower respiratory tract samples have increased plasma biomarkers of inflammation and worse outcomes. These findings suggest a possible mechanism of lung injury and inflammation in critical illness through interactions between the mycobiome and the innate immune system. Our study highlights the potential role of the largely understudied lung mycobiome as a contributor to clinical and biological heterogeneity in critically ill patients. Therefore, targeting the lung mycobiome may be a promising therapeutic approach for preventing and treating lung injury.

## Data Availability

Data Availability Statement: Sequencing data collected for this study are publicly available through the Sequencing Resource Archive, Accession number PRJNA726955

## Acknowledgments

The authors thank the patients and patient families that have enrolled in the cohort studies described in this report. They also thank the physicians, nurses, respiratory therapists, and other staff at the UPMC Presbyterian/Shadyside Hospitals for assistance with coordination of patient enrollment and collection of patient samples.

## IRB approval and informed consent

We enrolled subjects following admission to the hospital and obtained informed consent from the patients or their legally authorized representatives under the study protocol STUDY19050099 approved by the University of Pittsburgh Institutional Review Board (IRB).

## Data Availability Statement

Sequencing data collected for this study are publicly available through the Sequencing Resource Archive, Accession number PRJNA726955

## TABLES

**Table 2:** Baseline characteristics by Dirichlet Multinomial Model fungal clusters for endotracheal aspirates. Data are presented as median (with interquartile ranges) for continuous variables and n (%) for categorical variables. P-values for cluster comparisons were obtained from Wilcoxon test for continuous variables and Fisher’s test for categorical variables. Statistically significant p-values (p<0.05) are highlighted in bold.

|  | **Cluster 1**<br>**(N=136)** | **Cluster 2**<br>**(N=88)** | **p-value** |
| --- | --- | --- | --- |
| Age, median (IQR), years | 58.4 (43.8, 67.2) | 61.1 (52.0, 68.0) | 0.17 |
| Males, n (%) | 75 (55.1) | 50 (56.8) | 0.81 |
| BMI, median (IQR), kg/m<sup>2</sup> | 28.7 (25.2, 36.4) | 28.9 (24.8, 36.0) | 0.87 |
| Diabetes, n (%) | 48 (35.3) | 33 (37.5) | 0.74 |
| COPD, n (%) | 35 (25.7) | 22 (25.0) | 0.90 |
| Pulmonary fibrosis, n (%) | 8 (5.9) | 10 (11.4) | 0.14 |
| Immunosuppression, n (%) | 24 (17.6) | 27 (30.7) | 0.02 |
| ARDS, n (%) | 39 (28.7) | 17 (19.3) | 0.11 |
| Pneumonia, n (%) | 49 (36.0) | 37 (42.0) | 0.37 |
| Extrapulmonary sepsis, n (%) | 15 (11.0) | 18 (20.5) | 0.05 |
| Aspiration, n (%) | 20 (14.7) | 15 (17.0) | 0.64 |
| LIPS score, median (IQR) | 5.0 (4.0, 6.5) | 5.5 (4.0, 7.0) | 0.39 |
| SOFA score, median (IQR)* | 6.0 (4.0, 8.0) | 6.0 (4.0, 10.0) | 0.12 |
| PaO<sub>2</sub>/FiO<sub>2</sub> ratio, median (IQR), mm Hg | 164.0 (119.5, 205.0) | 166.0 (109.5, 233.2) | 0.70 |
| WBC, median (IQR), $\times 10^9$ cells/L | 11.4 (9.0, 16.2) | 12.9 (8.5, 16.9) | 0.57 |
| Plateau pressure, median (IQR), cm H<sub>2</sub>O | 20.0 (16.2, 25.0) | 19.0 (16.0, 25.0) | 0.71 |
| Positive respiratory cultures†, n (%) | 25 (19.1) | 22 (25.9) | 0.24 |
| Antibiotics before ICU admission, n (%) | 58 (42.6) | 29 (33.0) | 0.15 |
| Antibiotics during ICU admission before sampling, n (%) | 115 (84.6) | 75 (85.2) | 0.89 |
| Hyperinflammatory subphenotype, n (%) | 23 (17.4) | 20 (23.3) | 0.29 |

**Abbreviations:** IQR: interquartile range; BMI: body mass index; COPD: chronic obstructive pulmonary disease, LIPS: lung injury prediction score; SOFA: sequential organ failure assessment; PaO<sub>2</sub>: partial pressure of arterial oxygen; FiO<sub>2</sub>: Fractional inhaled concentration of oxygen; WBC: white blood cell count. \*SOFA score calculation does not include the neurologic component of SOFA score because all patients were intubated and receiving sedative medications, impairing our ability to perform an assessment of the Glasgow Coma Scale in a consistent and reproducible fashion. †Respiratory specimen cultures were defined as positive when yeast or fungal species were isolated by the clinical laboratory. Immunosuppression was defined as the receipt (up until admission) of chronic steroids, biologics, other immunosuppressive medications, cancer chemotherapy, or diagnosis of immunodeficiency.

