## Supplement for "Respiratory Fungal Communities are Associated with Systemic Inflammation and Predict Survival in Patients with Acute Respiratory Failure"

### METHODS:

#### Patient Enrollment:

We studied patients enrolled in the Acute Lung Injury Registry (ALIR) and Biospecimen Repository, an ongoing prospective observational cohort of critically ill patients at UPMC Presbyterian Hospital in Pittsburgh, Pennsylvania. From April 2015– September 2019, we prospectively enrolled adult patients with acute respiratory failure who were intubated and mechanically-ventilated in the Medical or Cardiac Intensive Care Units (ICU). Participants were enrolled within 72 hours of initiation of mechanical ventilation. Patients were excluded based on the following criteria: i) presence of tracheostomy, ii) expected survival <48hrs, and iii) inability to obtain informed consent. The study was approved by the University of Pittsburgh Institutional Review Board (STUDY19050099), and written informed consent was provided by all participants or their legally authorized representatives.

#### *Clinical Characteristics*

We extracted clinical data from the electronic medical record, including demographics, comorbid conditions, vital signs, laboratory results, and mechanical ventilation parameters within 24hrs from enrollment . We determined severity of illness by calculating the modified sequential organ failure assessment (SOFA) score (28), and lung injury risk with the Lung Injury Prediction Score (LIPS) (29). We recorded whether patients had received antibiotics in the 30 days prior to ICU admission and modeled the dosing duration, timing of administration, and specific type of systemic antibiotic exposure in the ICU (before microbiome sampling) as previously described.(30) We followed patients prospectively for time to liberation from mechanical ventilation and 30 day survival.(31)

#### *Clinical Sample Collection*

We sampled upper and lower respiratory tract mycobiota with a posterior oropharyngeal swab (oral) and an endotracheal aspirate (ETA), respectively, as previously described. (1) We collected blood samples for separation of plasma for measurement of host response biomarkers.

Distal tracheal secretions were suctioned through a closed endotracheal tube suctioning system (in-line suction catheter). In cases that suctioning did not return adequate (≥5ml) amount of ETAs, we instilled 5cc of sterile saline through the tubing system and then repeated suctioning. We also collected the left-over saline not used for sample collection (~ 5cc) as a negative control to examine for procedural contamination of our samples. Samples were directly collected in sputum collection traps, labeled and frozen to -80°C as quickly as possible and stored at this temperature until sample processing.

At study enrollment, we also collected blood samples (10 cc) (through central venous or arterial access or phlebotomy) and performed same-day centrifugation and plasma collection in the research laboratory and stored samples at -80°C.

#### *Clinical Microbiology*

We utilized clinical microbiologic results obtained within 48hrs of research biospecimens for the clinical microbiologic results to be of the same infectious process being studied by the next-generation sequencing. All clinical cultures were obtained at the discretion of the treating physicians and were not obtained specifically for research purposes. All specimen cultures were performed by the clinical microbiology laboratory at UPMC. Samples were used to inoculate agar plates: trypticase soy agar with 5% sheep blood, chocolate agar (for fastidious organisms), Columbia Naladixic Acid *Agar* (*CAN Agar for gram-positives)*, and MacConkey (for gram-negatives). Samples are incubated for up to 24hrs and the following day, isolated colonies are identified with matrix-assisted laser desorption/ionization with time-of-flight mass spectroscopy (MALDI-TOF MS) (Bruker Biotyper) and susceptibility testing is performed with the MicroScan WalkAway platform (Beckman Coulter).

#### *Host Response Biomarker Measurements*

We measured validated biomarkers of ARDS in plasma samples using a custom Luminex multi-analyte panel (R&D Systems, Minneapolis, MI, United States) for our host-response experiments. We included biomarkers targeting the innate immune response (IL-6, IL-8, IL-10, Tumor necrosis factor receptor 1 (TNFR1), suppression of tumorigenicity-2 [ST-2], fractalkine), *epithelial injury* (receptor of advanced glycation end-products [RAGE]), *endothelial injury* (Angiopoietin-2) and *host-response to infection* (procalcitonin and pentraxin-3) to assess host response in relationship to microbial profiles.(21–23) To assess for circulating fragments of fungal cell wall, we measured (1→3)-β-D-Glucan (BDG) with the Fungitell assay (Associates of Cape Cod, Inc.) in plasma samples.(38, 39) We defined a positive BDG test as greater or equal to the conventional threshold of 60 pg/ml.(40)

***DNA Isolation***

We extracted microbial DNA directly from ETA samples using the Powersoil (MoBio) extraction kit following the manufacturer's instructions, as previously described.(18) Due to the high viscosity of ETA samples, we pretreated them with Dithiothreitol (0.1% DTT in phosphate-buffered saline) in 1:1 dilution to dissolve the mucus and allow usability in DNA extraction columns.

***Fungal Gene Amplification and Sequencing***

We amplified extracted DNA by PCR using the method of Caporaso et al and the Q5 HS High-Fidelity polymerase (NEB) targeting the internal transcribed regions 1 and 2 of the ITS rRNA gene.(23, 24) We utilized reagent controls for each step of the process (DNA extraction and PCR amplification). We amplified four microliters per reaction of each sample with a single barcode in triplicate 25 microliter reactions. We utilized a 2-step nested PCR protocol. Initial cycle conditions were 98°C for 30 seconds, 15 cycles of 98°C for 10 seconds, 55°C for 30 seconds, and 72°C for 90 seconds followed by 72°C for 2 minutes. 5 microliters of the initial PCR reaction were added to the nested cycle and were processed using the following conditions: 98°C for 30 seconds, 15 cycles of 98°C for 10 seconds, 55°C for 30 seconds, and 72°C for 1 minute followed by 72°C for 2 minutes. We combined triplicates and purified with the AMPure XP beads (Beckman) at a 0.7:1 ratio (beads:DNA) to remove primer-dimers. We performed sample pooling on ice by combining 30 microliters of each sample. We purified the sample pool with the MinElute PCR purification kit. The final sample pool underwent two more purifications – AMPure XP beads to 0.7:1 to remove all traces of primer dimers and a final cleanup using the Purelink PCR Purification Kit (Life Technologies). We quantitated the purified pool in triplicate on the Qubit fluorimeter prior to preparing for sequencing. The sequencing pool was prepared according to instructions by Illumina (San Diego, CA), with an added incubation at 95˚C for 2 minutes immediately following the initial dilution to 20 picomolar.(22) We then diluted the sequencing pool to a final concentration of 7 pM + 15% PhiX control. Amplicons were sequenced on the Illumina Miseq platform. We collected experimental negative control samples used to assess for possible contamination events from sample collection to DNA extraction and PCR amplification.

***Experimental Controls***

Low biomass studies of the microbiome are susceptible to contamination by microbial DNA in the sample collection process and in reagents used in DNA extraction and amplification. We considered three types of experimental negative control samples used to assess for possible contamination events from sample collection to DNA extraction and PCR amplification: endotracheal aspirate collection controls, DNA extraction, and PCR amplification negative controls. ETA controls consisted of left-over sterile saline not used for sample collection as a negative control to identify potential contamination of the ETA sample from microbial populations present in clinically sterile saline syringes. We included these ETA controls in DNA extraction, PCR amplification, and sequencing alongside clinical samples. DNA extraction negative controls were comprised of sterile water in one DNA extraction column per batch of clinical samples undergoing DNA extraction. We added sterile water in the place of template DNA in our PCR reaction mix as negative PCR amplification controls. We also included PCR amplification positive controls to confirm effective amplification. We used the ZymoBIOMICS Microbial Community DNA Standard (Zymo Research, Irvine, CA), a mock microbial community consisting of genomic DNA of both bacteria and fungi as our PCR amplification positive control. Given the risk of contamination, we compared the fungal reads identified in clinical ETA samples that were distinct from negative experimental control samples to assess the amount of contamination impacting our sequencing results.

***ITS Sequence Processing***

ITS rRNA sequences from the pooled sequencing run were demultiplexed into individual sample/replicate fastq files. The variable-length reads were processed, trimmed and quality filtered with a quality control pipeline utilizing the DADA2 package in R.(21) Paired sequences with forward and reverse reads passing the quality filtering and trimming steps were merged. Chimeras were removed and the Unite database will be utilized to classify reads into amplicon sequence variants (ASVs) using the naïve Bayesian classifier method.(22, 23) ASVs are inferred based on sequence data and can be defined at different levels of resolution (phylum, class, order, family, genus, and species). Results of ASV annotation were converted into per-sample taxonomic profiles represented as categorical counts in matrices of dimension (number of samples) x number of categories). The taxa table was filtered for low abundance taxa (relative abundance, <0.005%) and singletons, and for samples that generated fewer than 100 reads in the range of negative control samples.

**RESULTS:**

**Supplementary Table 1:** Baseline and Clinical Characteristics of All Patients with Samples Sequenced

| | **Overall (n=316)** |
|---|---|
| Age, median (IQR), years | 59.0 (45.7, 68.0) |
| Males, n (%) | 172.0 (54.4%) |
| BMI, median (IQR), kg/m2 | 29.0 (25.4, 35.7) |
| Diabetes, n (%) | 113.0 (35.8%) |
| COPD, n (%) | 76.0 (24.1%) |
| Pulmonary fibrosis, n (%) | 20.0 (6.3%) |
| Immunosuppression, n (%) | 67.0 (21.2%) |
| ARDS, n (%) | 79.0 (25.0%) |
| Pneumonia, n (%) | 121.0 (38.3%) |
| Extrapulmonary sepsis, n (%) | 50.0 (15.8%) |
| Aspiration, n (%) | 56.0 (17.7%) |
| LIPS score, median (IQR) | 5.2 (4.0, 6.5) |
| SOFA score, median (IQR)* | 6.0 (4.0, 8.0) |
| PaO2/FiO2ratio, median (IQR), mm Hg | 166.5 (118.0, 206.5) |
| WBC, median (IQR), ×10^9cells/L | 11.7 (8.4, 16.7) |
| Plateau pressure, median (IQR), cm H2O | 20.0 (16.0, 25.0) |
| Positive respiratory cultures†, n (%) | 54.0 (18.9%) |
| Antibiotics before ICU admission, n (%) | 121.0 (38.3%) |
| Antibiotics during ICU admission before sampling, n (%) | 272.0 (86.1%) |
| Hyperinflammatory subphenotype, n (%) | 65.0 (21.3%) |

**Abbreviations:** IQR: interquartile range; BMI: body mass index; COPD: chronic obstructive pulmonary disease, LIPS: lung injury prediction score; SOFA: sequential organ failure assessment; PaO2: partial pressure of arterial oxygen; FiO2: Fractional inhaled concentration of oxygen; WBC: white blood cell count. *SOFA score calculation does not include the neurologic component of SOFA score because all patients were intubated and receiving sedative medications, impairing our ability to perform assessment of the Glasgow Coma Scale in a consistent and reproducible fashion. †Respiratory specimen cultures defined as positive when yeast or fungal species were isolated by the clinical laboratory.

**Supplementary Table 2:** Baseline and Clinical Characteristics of Patients with ETA Samples Sequenced

| | Filtered for Low Reads (N=78) | Included in Analysis (N=238) | p value |
|---|---|---|---|
| Age, median (IQR), years | 57.3 (45.3, 68.1) | 60.1 (46.4, 67.8) | 0.65 |
| Males, n (%) | 38.0 (48.7%) | 134.0 (56.3%) | 0.24 |
| BMI, median (IQR), kg/m2 | 29.3 (25.7, 34.5) | 28.8 (25.2, 36.2) | 0.70 |
| Diabetes, n (%) | 28.0 (35.9%) | 85.0 (35.7%) | 0.98 |
| COPD, n (%) | 19.0 (24.4%) | 57.0 (23.9%) | 0.94 |
| Pulmonary fibrosis, n (%) | 1.0 (1.3%) | 19.0 (8.0%) | **0.03** |
| Immunosuppression, n (%) | 11.0 (14.1%) | 56.0 (23.5%) | 0.08 |
| ARDS, n (%) | 19.0 (24.4%) | 60.0 (25.2%) | 0.88 |
| Pneumonia, n (%) | 30.0 (38.5%) | 91.0 (38.2%) | 0.97 |
| Extrapulmonary sepsis, n (%) | 14.0 (17.9%) | 36.0 (15.1%) | 0.55 |
| Aspiration, n (%) | 15.0 (19.2%) | 41.0 (17.2%) | 0.69 |
| LIPS score, median (IQR) | 5.0 (4.0, 6.5) | 5.5 (4.0, 7.0) | 0.83 |
| SOFA score, median (IQR)* | 5.0 (4.0, 8.0) | 6.0 (4.0, 8.0) | 0.27 |
| PaO2/FiO2ratio, median (IQR), mm Hg | 177.0 (122.5, 209.0) | 164.0 (117.2, 205.8) | 0.62 |
| WBC, median (IQR), ×10^9cells/L | 11.1 (7.9, 15.9) | 11.9 (8.9, 16.9) | 0.39 |
| Plateau pressure, median (IQR), cm H2O | 19.0 (15.0, 24.0) | 20.0 (16.0, 25.0) | 0.38 |
| Positive respiratory cultures†, n (%) | 5.0 (8.3%) | 49.0 (21.7%) | **0.02** |
| Antibiotics before ICU admission, n (%) | 27.0 (34.6%) | 94.0 (39.5%) | 0.44 |
| Antibiotics during ICU admission before sampling, n (%) | 68.0 (87.2%) | 204.0 (85.7%) | 0.75 |
| Hyperinflammatory subphenotype, n (%) | 15.0 (20.5%) | 50.0 (21.6%) | 0.86 |

**Abbreviations:** IQR: interquartile range; BMI: body mass index; COPD: chronic obstructive pulmonary disease, LIPS: lung injury prediction score; SOFA: sequential organ failure assessment; PaO2: partial pressure of arterial oxygen; FiO2: Fractional inhaled concentration of oxygen; WBC: white blood cell count. *SOFA score calculation does not include the neurologic component of SOFA score because all patients were intubated and receiving sedative medications, impairing our ability to perform assessment of the Glasgow Coma Scale in a consistent and reproducible fashion. †Respiratory specimen cultures defined as positive when yeast or fungal species were isolated by the clinical laboratory.

**Supplementary Table 3:** Baseline and Clinical Characteristics of Patients with Oral Samples Sequenced

| | Filtered for Low Reads (N=20) | Included in Analysis (N=202) | p value |
|---|---|---|---|
| Age, median (IQR), years | 59.9 (49.0, 68.0) | 60.4 (47.8, 68.5) | 0.90 |
| Males, n (%) | 16.0 (80.0%) | 103.0 (51.0%) | **0.01** |
| BMI, median (IQR), kg/m2 | 28.1 (24.2, 33.0) | 28.3 (25.2, 36.0) | 0.46 |
| Diabetes, n (%) | 8.0 (40.0%) | 65.0 (32.2%) | 0.48 |
| COPD, n (%) | 6.0 (30.0%) | 45.0 (22.3%) | 0.43 |
| Pulmonary fibrosis, n (%) | 2.0 (10.0%) | 11.0 (5.4%) | 0.41 |
| Immunosuppression, n (%) | 4.0 (20.0%) | 45.0 (22.3%) | 0.81 |
| ARDS, n (%) | 6.0 (30.0%) | 34.0 (16.8%) | 0.14 |
| Pneumonia, n (%) | 10.0 (50.0%) | 70.0 (34.7%) | 0.17 |
| Extrapulmonary sepsis, n (%) | 2.0 (10.0%) | 28.0 (13.9%) | 0.63 |
| Aspiration, n (%) | 6.0 (30.0%) | 33.0 (16.3%) | 0.13 |
| LIPS score, median (IQR) | 5.5 (4.2, 6.2) | 5.0 (3.9, 6.5) | 0.85 |
| SOFA score, median (IQR)* | 5.0 (4.0, 5.8) | 6.0 (4.0, 8.0) | 0.08 |
| PaO2/FiO2ratio, median (IQR), mm Hg | 145.5 (133.2, 205.0) | 180.5 (127.0, 219.5) | 0.36 |
| WBC, median (IQR), ×10^9cells/L | 8.6 (6.9, 14.7) | 11.4 (8.3, 16.6) | 0.13 |
| Plateau pressure, median (IQR), cm H2O | 19.5 (14.0, 23.2) | 19.0 (16.0, 24.0) | 0.84 |
| Positive respiratory cultures†, n (%) | 3.0 (15.8%) | 33.0 (17.8%) | 0.82 |
| Antibiotics before ICU admission, n (%) | 8.0 (40.0%) | 89.0 (44.1%) | 0.73 |
| Antibiotics during ICU admission before sampling, n (%) | 18.0 (90.0%) | 174.0 (86.1%) | 0.63 |
| Hyperinflammatory subphenotype, n (%) | 0.0 (0.0%) | 38.0 (19.3%) | **0.04** |

**Abbreviations:** IQR: interquartile range; BMI: body mass index; COPD: chronic obstructive pulmonary disease, LIPS: lung injury prediction score; SOFA: sequential organ failure assessment; PaO2: partial pressure of arterial oxygen; FiO2: Fractional inhaled concentration of oxygen; WBC: white blood cell count. *SOFA score calculation does not include the neurologic component of SOFA score because all patients were intubated and receiving sedative medications, impairing our ability to perform assessment of the Glasgow Coma Scale in a consistent and reproducible fashion. †Respiratory specimen cultures defined as positive when yeast or fungal species were isolated by the clinical laboratory.

**Supplementary Table 4:** Clinical Outcomes by ETA DMM Cluster and Samples Filtered for Low Reads

| | **Unadjusted** | | | **Adjusted** | | |
|---|---|---|---|---|---|---|
| | HR | 95% CI | P value | HR | 95% CI | P value |
| **30 Day Mortality** | | | | | | |
| ETA DMM Cluster 2 | 2.2 | 1.30, 3.73 | **0.003** | 1.83 | 1.04, 3.21 | **0.036** |
| Filtered for Low Reads | 0.69 | 0.33, 1.44 | 0.33 | 0.84 | 0.37, 1.90 | 0.68 |

**Supplementary Table 5:** ITS Sequencing Reads by Sample Type in All Samples Sequenced – Comparisons reported against ETA sample number of reads (surrogate for fungal load).

| Sample Type | Fungal Sequencing Reads | p-value |
|---|---|---|
| ETA (N=331) | 3288.0 (109.5, 27245.0) | |
| Oral (N=256) | 20828.5 (2367.5, 49991.0) | **<0.0001** |
| DNA Extraction Negative (N=8) | 1.0 (1.0, 1.0) | **<0.0001** |
| DNA Extraction Positive (N=34) | 2156.0 (138.2, 3369.2) | **0.04** |
| PCR Negative (N=26) | 1.0 (0.0, 1.8) | **<0.0001** |
| PCR Positive (N=28) | 49442.0 (34488.0, 74675.5) | **<0.0001** |

**Abbreviations:** DNA: Deoxyribonucleic acid; ETA: endotracheal aspirate; PCR: polymerase chain reaction

**Supplementary Table 6:** Mycobiome Characteristics by Sample Type in Samples Included in Analysis

| | **ETA (N=238)** | **Oral (N=202)** | **p value** |
|---|---|---|---|
| Fungal sequencing reads, median (IQR) | 10029.50 (1115.75, 42466.50) | 26039.00 (3775.00, 55997.00) | **< 0.01** |
| Shannon Index, median (IQR) | 0.35 (0.11, 0.81) | 0.46 (0.11, 0.89) | 0.19 |
| Mycobiome Composition, n(%) | | | |
| *Multiple Fungi* | 170.0 (71.4%) | 165.0 (81.7%) | |
| *Monofungal* | 68.0 (28.6%) | 37.0 (18.3%) | **0.01** |

**Supplementary Table 7:** Biomarkers by Fungal DMM Cluster in ETA samples

| | Cluster 1 (N=136) | Cluster 2 (N=88) | p value |
|---|---|---|---|
| IL-6 (log) median (IQR) | 3.63 (2.68, 4.98) | 3.84 (3.17, 5.09) | 0.20 |
| IL-8 (log) median (IQR) | 2.58 (1.91, 3.23) | 2.95 (2.32, 3.49) | **0.01** |
| IL-10 (log) median (IQR) | 0.80 (-0.24, 2.13) | 1.18 (-0.24, 2.41) | 0.33 |
| TNFr1 (log) median (IQR) | 7.95 (7.26, 8.71) | 8.01 (7.60, 9.02) | 0.17 |
| ST-2 (log) median (IQR) | 11.80 (10.89, 12.77) | 12.10 (11.36, 12.84) | 0.09 |
| Fractalkine (log) median (IQR) | 7.35 (6.60, 7.81) | 7.46 (6.89, 7.91) | 0.32 |
| RAGE (log) median (IQR) | 7.90 (7.42, 8.59) | 7.95 (7.42, 8.50) | 0.90 |
| ANG-2 (log) median (IQR) | 8.54 (7.87, 9.29) | 8.97 (8.05, 9.71) | 0.06 |
| Pentraxin-3 (log) median (IQR) | 7.80 (6.79, 8.85) | 8.32 (7.51, 9.25) | **0.02** |
| Procalcitonin (log) median (IQR) | 6.09 (4.84, 7.89) | 6.22 (5.13, 7.74) | 0.51 |
| Plasma 1,3-β-d-glucan (log) median (IQR) | 3.24 (2.71, 3.83) | 3.31 (2.77, 3.81) | 0.93 |
| Plasma 1,3-β-d-glucan Greater than 60 pg/mL, n (%) | 22.0 (18.8%) | 13.0 (17.6%) | 0.83 |

**Abbreviations:** IL-1β = interleukin 1 beta; IL-3 = interleukin 3; IL-6 = interleukin 6; IL-8 = interleukin 8; IL-10 = interleukin 10; IFN-γ= interferon gamma; TNFr1= tumor necrosis factor receptor 1; ST-2= suppression of tumorigenicity 2; RAGE= receptor of advanced glycation end products; ANG-2 = angiopoietin 2; SPD = surfactant protein D.

**Supplementary Table 8:** Linear Regression of Biomarkers by Fungal DMM Cluster in ETA samples

| ETA DMM Cluster 2 | | | | | | |
|---|---|---|---|---|---|---|
| | Unadjusted | | | Adjusted | | |
| | Beta | 95% CI | p value | Beta | 95% CI | p value |
| IL-6 (log) | 0.03 | -0.01, 0.07 | 0.17 | 0.03 | -0.01, 0.07 | 0.10 |
| IL-8 (log) | 0.06 | 0.01, 0.12 | **0.03** | 0.06 | 0.01, 0.12 | **0.03** |
| IL-10 (log) | 0.02 | -0.02, 0.06 | 0.33 | 0.03 | -0.01, 0.07 | 0.14 |
| TNFr1 (log) | 0.05 | -0.04, 0.15 | 0.25 | 0.04 | -0.06, 0.14 | 0.44 |
| ST-2 (log) | 0.04 | -0.01, 0.10 | 0.12 | 0.03 | -0.02, 0.09 | 0.22 |
| Fractalkine (log) | 0.04 | -0.03, 0.12 | 0.27 | 0.05 | -0.03, 0.13 | 0.20 |
| RAGE (log) | 0.01 | -0.06, 0.08 | 0.82 | 0.02 | -0.05, 0.09 | 0.61 |
| ANG2 (log) | 0.05 | -0.01, 0.11 | 0.09 | 0.06 | 0.00, 0.12 | **0.04** |
| Pentraxin-3 (log) | 0.06 | 0.01, 0.10 | **0.009** | 0.05 | 0.01, 0.09 | **0.01** |
| Procalcitonin (log) | 0.01 | -0.03, 0.05 | 0.49 | 0.02 | -0.02, 0.06 | 0.33 |
| Plasma 1,3-β-d-glucan (log) | 0.00 | -0.07, 0.07 | 0.99 | 0.00 | -0.07, 0.07 | 0.96 |
| Plasma 1,3-β-d-glucan Greater than 60 pg/mL | 0.98 | 0.82, 1.17 | 0.83 | 0.96 | 0.80, 1.14 | 0.62 |

Estimates (Beta) with corresponding 95% confidence intervals were derived from the multivariate linear models adjusted for age, SOFA score, history of pulmonary fibrosis, and positive respiratory fungal culture.
**Abbreviations:** IL-1β = interleukin 1 beta; IL-3 = interleukin 3; IL-6 = interleukin 6; IL-8 = interleukin 8; IL-10 = interleukin 10; IFN-γ= interferon gamma; TNFr1= tumor necrosis factor receptor 1; ST-2= suppression of tumorigenicity 2; RAGE= receptor of advanced glycation end products; ANG-2 = angiopoietin 2

**Supplementary Table 9:** Biomarkers by Respiratory Fungal Culture Status

| | Negative (N=177) | Positive (N=49) | p value |
|---|---|---|---|

| | | | |
|---|---|---|---|
| IL-6 (log) median (IQR) | 3.64 (2.89, 4.99) | 4.68 (3.49, 5.29) | **0.01** |
| IL-8 (log) median (IQR) | 2.67 (1.99, 3.30) | 2.97 (2.44, 3.64) | **0.04** |
| IL-10 (log) median (IQR) | 0.80 (-0.24, 2.21) | 0.70 (-0.24, 2.34) | 0.62 |
| TNFr1 (log) median (IQR) | 7.89 (7.26, 8.71) | 8.58 (7.91, 9.14) | **< 0.01** |
| ST-2 (log) median (IQR) | 11.87 (11.14, 12.88) | 12.09 (11.10, 12.49) | 0.78 |
| Fractalkine (log) median (IQR) | 7.36 (6.61, 7.88) | 7.46 (6.97, 7.86) | 0.46 |
| RAGE (log) median (IQR) | 7.90 (7.42, 8.44) | 8.38 (7.71, 9.06) | **< 0.01** |
| ANG-2 (log) median (IQR) | 8.73 (7.91, 9.46) | 8.61 (8.20, 9.53) | 0.60 |
| Pentraxin-3 (log) median (IQR) | 7.86 (6.96, 8.88) | 8.65 (7.52, 9.30) | 0.10 |
| Procalcitonin (log) median (IQR) | 6.21 (4.95, 7.65) | 6.21 (5.26, 8.00) | 0.36 |
| Plasma 1,3-β-d-glucan (log) median (IQR) | 3.22 (2.71, 3.66) | 3.53 (2.89, 4.45) | 0.04 |
| Plasma 1,3-β-d-glucan Greater than 60 pg/mL, n (%) | 23.0 (14.6%) | 11.0 (26.2%) | 0.07 |

**Abbreviations:** IL-1β = interleukin 1 beta; IL-3 = interleukin 3; IL-6 = interleukin 6; IL-8 = interleukin 8; IL-10 = interleukin 10; IFN-γ= interferon gamma; TNFr1= tumor necrosis factor receptor 1; ST-2= suppression of tumorigenicity 2; RAGE= receptor of advanced glycation end products; ANG-2 = angiopoietin 2

**Supplementary Table 10:** Linear Regression of Biomarkers by Respiratory Fungal Culture Status

| **Positive Respiratory Fungal Culture Status** | | | | | | |
|---|---|---|---|---|---|---|
| | **Unadjusted** | | | **Adjusted** | | |
| | Beta | 95% CI | p value | Beta | 95% CI | p value |
| IL-6 (log) | 0.04 | 0.01, 0.07 | **0.02** | 0.05 | 0.01, 0.09 | **0.007** |
| IL-8 (log) | 0.05 | 0.00, 0.09 | **0.04** | 0.06 | 0.00, 0.11 | **0.03** |
| IL-10 (log) | 0.01 | -0.02, 0.04 | 0.60 | 0.02 | -0.02, 0.06 | 0.27 |
| TNFr1 (log) | 0.12 | 0.05, 0.19 | **0.002** | 0.20 | 0.10, 0.30 | **<0.001** |
| ST-2 (log) | -0.01 | -0.05, 0.04 | 0.80 | 0.00 | -0.05, 0.05 | 0.96 |
| Fractalkine (log) | 0.03 | -0.04, 0.09 | 0.38 | 0.05 | -0.02, 0.12 | 0.19 |
| RAGE (log) | 0.08 | 0.03, 0.14 | **0.004** | 0.11 | 0.05, 0.17 | **<0.001** |
| ANG2 (log) | 0.02 | -0.03, 0.07 | 0.43 | 0.05 | -0.02, 0.11 | 0.17 |
| Pentraxin-3 (log) | 0.03 | -0.01, 0.06 | 0.12 | 0.04 | 0.00, 0.08 | **0.05** |
| Procalcitonin (log) | 0.02 | -0.02, 0.05 | 0.29 | 0.03 | -0.01, 0.07 | 0.14 |
| Plasma 1,3-β-d-glucan (log) | 0.08 | 0.02, 0.14 | **0.01** | 0.09 | 0.03, 0.15 | **0.005** |
| Plasma 1,3-β-d-glucan Greater than 60 pg/mL | 0.14 | -0.01, 0.29 | 0.07 | 0.13 | -0.03, 0.29 | 0.11 |

Estimates (Beta) with corresponding 95% confidence intervals were derived from the multivariate linear models adjusted for age, SOFA score and history of pulmonary fibrosis. **Abbreviations:** IL-1β = interleukin 1 beta; IL-3 = interleukin 3; IL-6 = interleukin 6; IL-8 = interleukin 8; IL-10 = interleukin 10; IFN-γ= interferon gamma; TNFr1= tumor necrosis factor receptor 1; ST-2= suppression of tumorigenicity 2; RAGE= receptor of advanced glycation end products; ANG-2 = angiopoietin 2

**Supplementary Table 11:** Biomarkers by C.albicans Relative Abundance

| | **C.albicans ≥ 50% (N=121)** | **C.albicans <50% (N=117)** | **p value** |
|---|---|---|---|
| IL-6 (log) median (IQR) | 3.83 (2.94, 5.13) | 3.79 (2.99, 5.02) | 0.89 |
| IL-8 (log) median (IQR) | 2.88 (2.09, 3.49) | 2.64 (1.98, 3.26) | 0.22 |
| IL-10 (log) median (IQR) | 0.80 (-0.24, 2.41) | 0.47 (-0.24, 2.08) | 0.26 |
| TNFr1 (log) median (IQR) | 8.04 (7.56, 8.83) | 7.96 (7.24, 8.80) | 0.29 |
| ST-2 (log) median (IQR) | 12.09 (11.22, 12.90) | 11.80 (11.00, 12.73) | 0.20 |
| Fractalkine (log) median (IQR) | 7.52 (6.77, 7.93) | 7.28 (6.64, 7.81) | 0.29 |

| | | | |
|---|---|---|---|
| RAGE (log) median (IQR) | 7.95 (7.44, 8.58) | 7.99 (7.43, 8.60) | 0.93 |
| ANG-2 (log) median (IQR) | 8.90 (8.05, 9.65) | 8.63 (7.91, 9.42) | 0.21 |
| Pentraxin-3 (log) median (IQR) | 8.12 (7.38, 9.22) | 7.95 (6.85, 8.88) | 0.12 |
| Procalcitonin (log) median (IQR) | 6.26 (5.09, 7.93) | 6.38 (5.11, 7.93) | 0.98 |
| Plasma 1,3-β-d-glucan (log) median (IQR) | 3.31 (2.77, 3.74) | 3.22 (2.71, 3.96) | 0.82 |
| Plasma 1,3-β-d-glucan Greater than 60 pg/mL, n (%) | 16.0 (15.4%) | 21.0 (20.8%) | 0.31 |

**Abbreviations:** IL-1β = interleukin 1 beta; IL-3 = interleukin 3; IL-6 = interleukin 6; IL-8 = interleukin 8; IL-10 = interleukin 10; IFN-γ= interferon gamma; TNFr1= tumor necrosis factor receptor 1; ST-2= suppression of tumorigenicity 2; RAGE= receptor of advanced glycation end products; ANG-2 = angiopoietin 2

**Supplementary Table 12:** Linear Regression of Biomarkers by C.albicans Relative Abundance

| **C.albicans >50% Relative Abundance** | | | | | | |
|---|---|---|---|---|---|---|
| | **Unadjusted** | | | **Adjusted** | | |
| | Beta | 95% CI | p value | Beta | 95% CI | p value |
| IL-6 (log) | 0.00 | -0.03, 0.04 | 0.95 | 0.00 | -0.04, 0.04 | 0.93 |
| IL-8 (log) | 0.01 | -0.04, 0.06 | 0.59 | 0.02 | -0.04, 0.08 | 0.52 |
| IL-10 (log) | 0.02 | -0.02, 0.06 | 0.33 | 0.04 | -0.01, 0.08 | 0.11 |
| TNFr1 (log) | 0.05 | -0.05, 0.14 | 0.32 | 0.12 | -0.01, 0.25 | 0.07 |
| ST-2 (log) | 0.03 | -0.02, 0.08 | 0.29 | 0.04 | -0.02, 0.10 | 0.17 |
| Fractalkine (log) | 0.04 | -0.04, 0.11 | 0.31 | 0.03 | -0.05, 0.12 | 0.43 |
| RAGE (log) | 0.02 | -0.05, 0.09 | 0.58 | 0.03 | -0.05, 0.11 | 0.45 |
| ANG-2 (log) | 0.03 | -0.03, 0.09 | 0.26 | 0.07 | -0.01, 0.14 | 0.10 |
| Pentraxin-3 (log) | 0.04 | 0.00, 0.08 | **0.05** | 0.04 | 0.00, 0.09 | 0.06 |
| Procalcitonin (log) | 0.00 | -0.04, 0.04 | 0.94 | 0.01 | -0.04, 0.06 | 0.66 |
| Plasma 1,3-β-d-glucan (log) | 0.00 | -0.07, 0.07 | 0.98 | 0.00 | -0.08, 0.07 | 0.92 |
| Plasma 1,3-β-d-glucan Greater than 60 pg/mL | -0.09 | -0.27, 0.09 | 0.32 | -0.13 | -0.32, 0.06 | 0.16 |

**Abbreviations:** IL-1β = interleukin 1 beta; IL-3 = interleukin 3; IL-6 = interleukin 6; IL-8 = interleukin 8; IL-10 = interleukin 10; IFN-γ= interferon gamma; TNFr1= tumor necrosis factor receptor 1; ST-2= suppression of tumorigenicity 2; RAGE= receptor of advanced glycation end products; ANG-2 = angiopoietin 2

**FIGURES:**

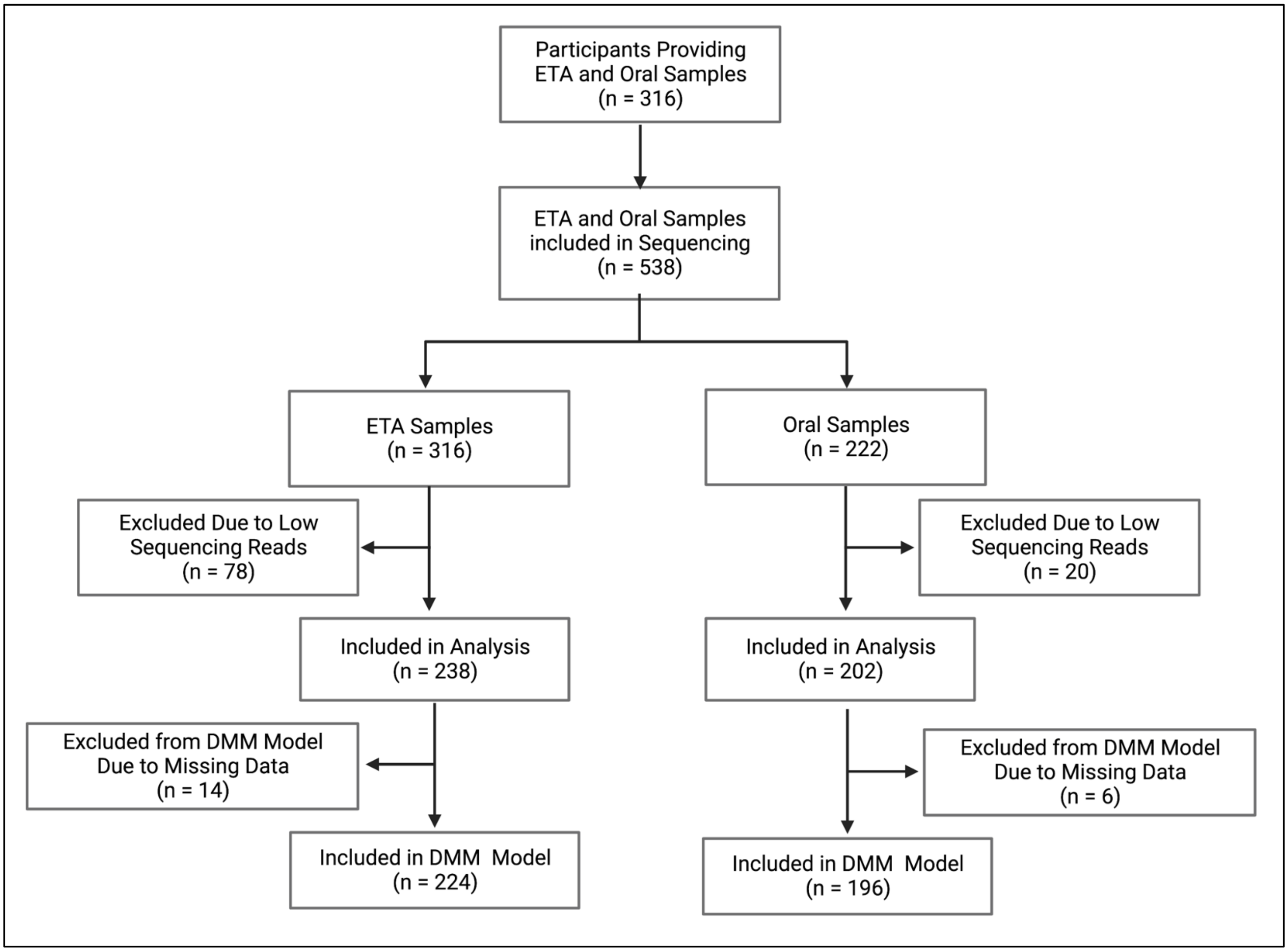


**Supplementary Figure 1:** Participant and sample inclusion.

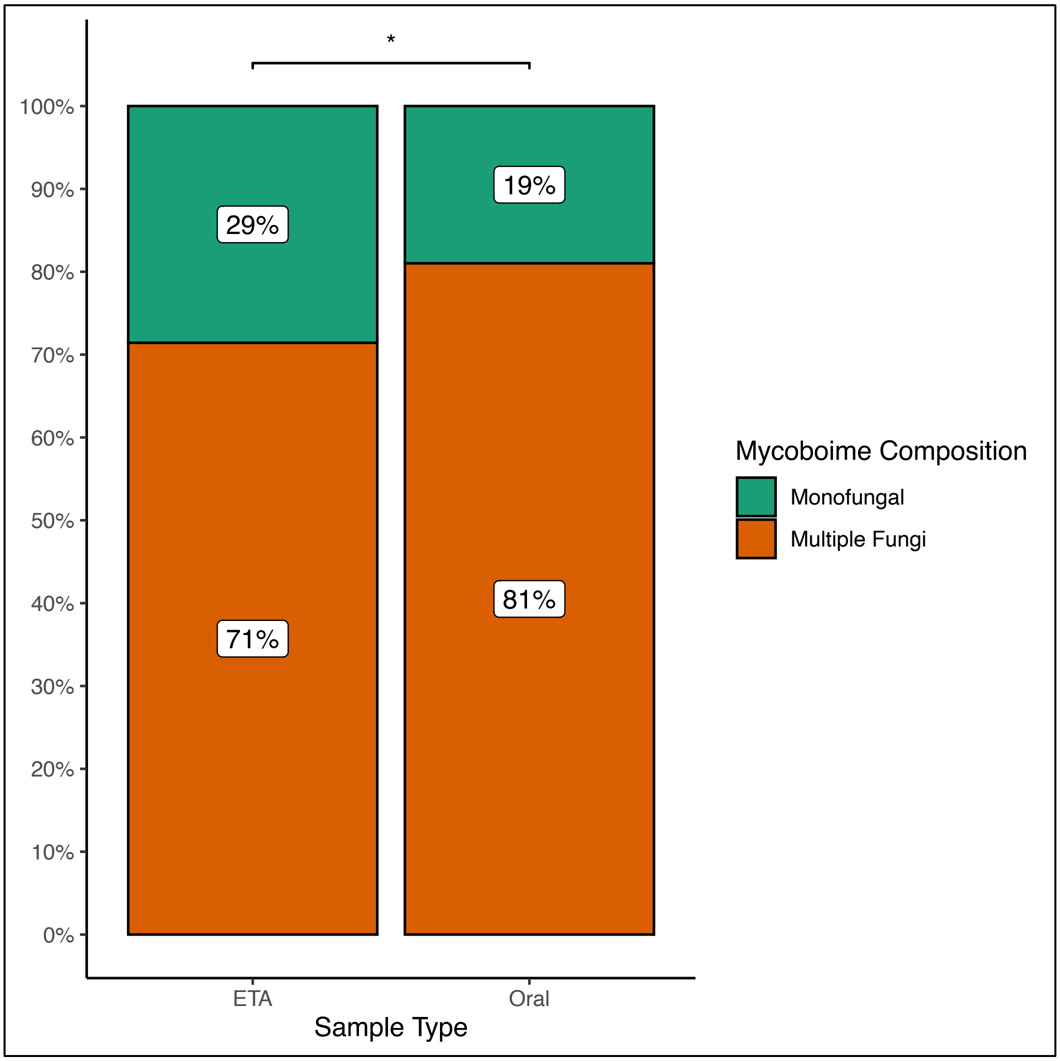


**Supplementary Figure 2:** Mycobiome composition by sample type. Fisher's Exact test (p=0.01).

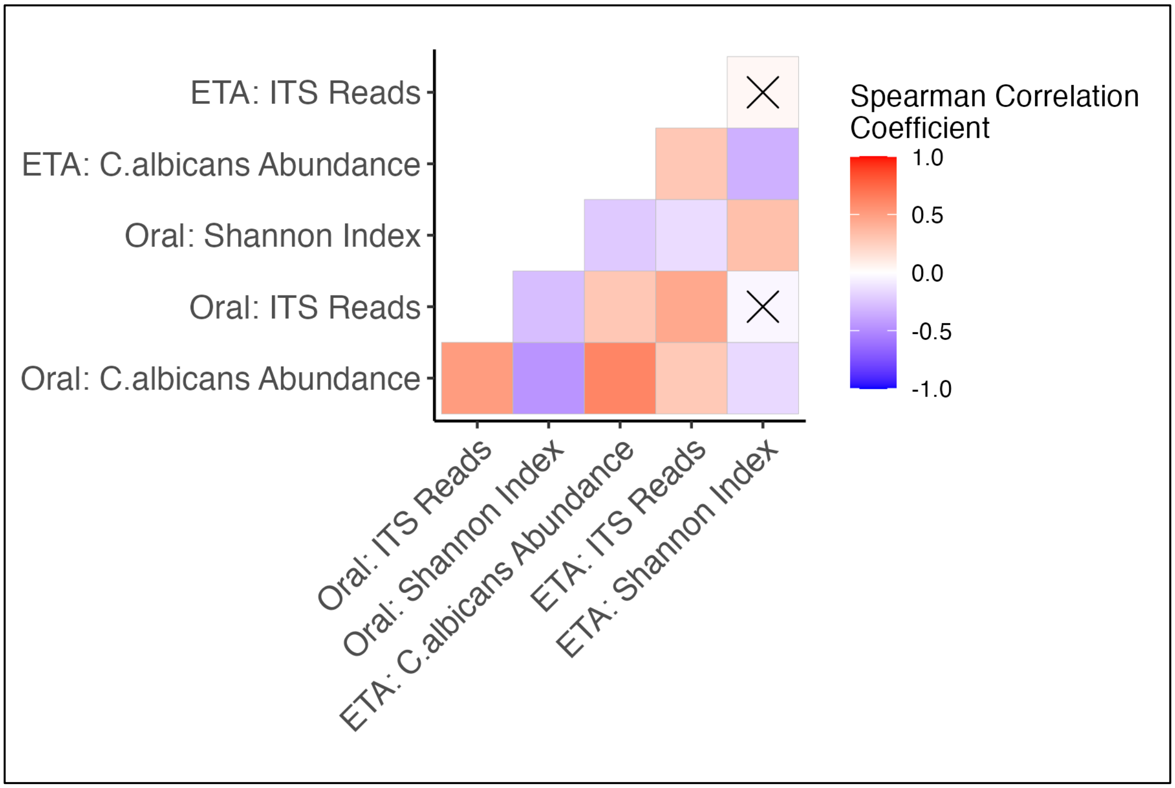


**Supplementary Figure 3:** Spearman Correlations of Mycobiome Characteristics Between Oral and ETA Samples. Correlations marked with an X indicate p-values >0.05.

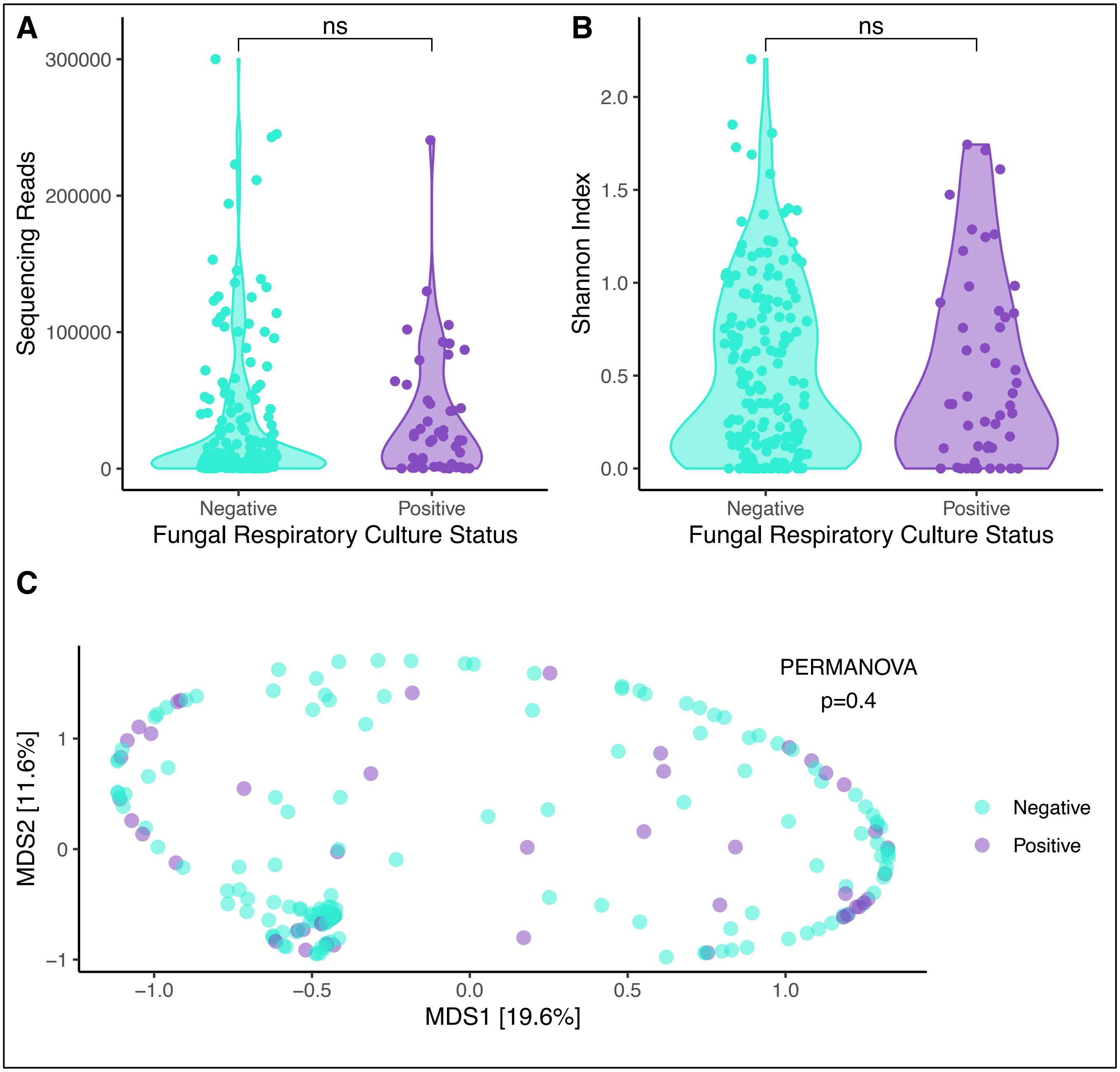


**Supplementary Figure 4: Fungal culture results did not explain differneces in fungal community profiles.** (A) Number of high quality ITS reads per sample are shown on the y-axis. No significant difference in the number of sequencing reads was observed between positive and negative respiratory cultures when yeast was considered to be a positive culture result. (B) The a-diversity comparisons between culture results showed no difference in the Shannon index. (C) Principal coordinates analyses for b-diversity comparisons (Manhattan distances) with Permanova for all samples included and stratified by culture results. ETA= endotracheal aspirate; Permanova = permutational ANOVA; MDS = multidimensional scaling axes; ns: p > 0.05; * p <= 0.05; ** p <= 0.01; *** p <= 0.001; **** p <= 0.0001

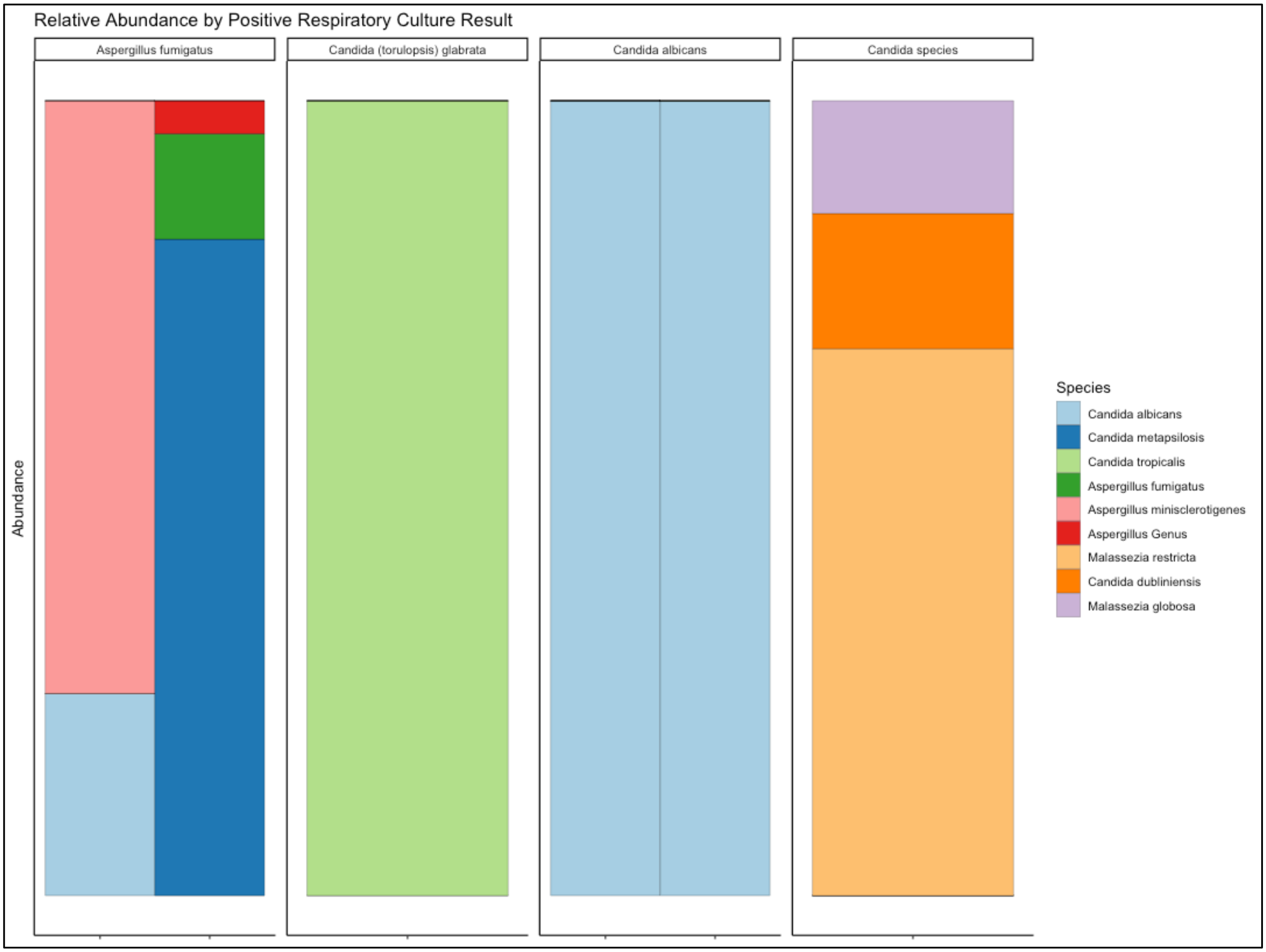


**Supplementary Figure 5: Taxonomic compositional differences by Positive Fungal Respiratory Fungal Culture Results.** Taxonomic bar plots for individual endotracheal aspirate samples, stratified by fungal species reported in positive respiratory fungal cultures. Taxonomic composition is shown as stacked bar graphs, with each bar representing the fungal community of each cluster, with taxa colored individually and widths of component bars corresponding to the relative abundance of each species.

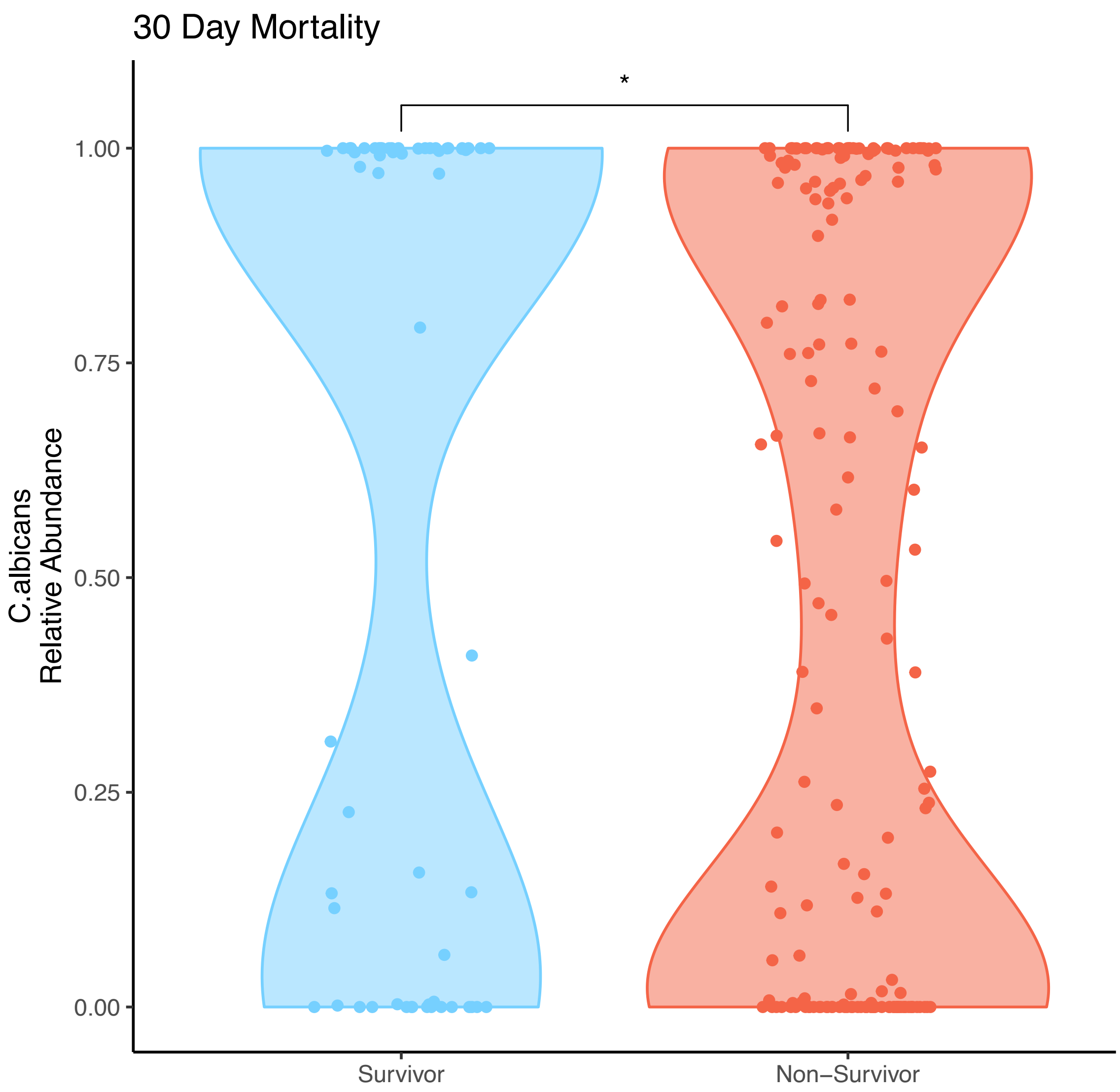


**Supplementary Figure 6:** C. albicans Relative Abundance by Mortality 30 Days from ICU Admission.
